# Repetitive Transcranial Magnetic Stimulation in Substance Use Disorders is Safe and Tolerable: A Systematic Review of 141 Clinical Trials Including 4,299 Participants

**DOI:** 10.1101/2025.10.02.25337151

**Authors:** Afra Souki, Shermin Sharifzadeh, Kathleen Brady, Colleen A Hanlon, Manish Kumar Jha, Kelvin O Lim, Gopalkumar Rakesh, Jonathan R Young, Vaughn R Steele, Victor M Tang, Hamed Ekhtiari

## Abstract

**Background:** Repetitive transcranial magnetic stimulation (rTMS) is a noninvasive neuromodulation intervention that has been investigated as a treatment for substance use disorder (SUD) and its co-occurring disorders. As the number of rTMS SUD clinical trials increase, the safety and tolerability profile should be assessed. In this systematic review, we investigate adverse events (AEs) of rTMS in individuals with SUD and factors that may influence their occurrence, including treatment status, and substance-use type.

**Methods:** We performed a systematic PubMed search to identify all controlled trials of rTMS in SUD published up to January 2025. Eligible studies were assessed, and safety information was extracted for analysis.

**Results:** A total of 141 controlled clinical trials with 4,299 participants were included in their active arms. rTMS trials recruited participants who were engaged in active substance use (37 studies), in the pre-treatment phase (61), in early recovery (18), or in sustained recovery (25). Twenty-two studies (15.60 %) explicitly reported no AEs adverse effects. Sixty-nine (48.94 %) studies reported only mild AEs adverse events, while only six studies (4.26%) reported moderate AEs adverse events. Thirty-five studies did not report safety-outcomes/AEs. As expected, participants reported mild and temporary AEs such as headaches, pain or discomfort under the coil, or dizziness. The most frequent side effects were headache (57 studies in active TMS vs. 32 in sham TMS), pain or discomfort at the stimulation site (34 vs. 20 studies), and dizziness (11 vs. 5 studies). These effects were generally temporary and did not require participants to stop treatment. Only nine studies reported serious side effects (7 studies in active TMS and 2 in sham TMS). Importantly, no seizures attributable to active rTMS were reported in these SUD samples. The other severe events included severe headache (5 studies), tinnitus (1), disorientation (1), and suicidal ideation (1). In total, 9 studies reported dropout due to adverse effects. Thirty-five studies (24.82%) did not report safety-outcomes/adverse-events.

**Conclusion:** Overall, rTMS in SUD samples is safe and well-tolerated regardless of recovery stage and substance. Most reported side effects were mild, self-limiting, and tolerable. This evidence supports the safety of rTMS as a potential stand-alone or adjunctive treatment for SUD.

**Highlights:**

- We reviewed 141 controlled rTMS trials with 4,299 SUD participants
- rTMS in SUDs is safe and well-tolerated
- No reports of rTMS-induced seizures among 4,299 participants
- Most side-effects were mild and transient (headache, scalp pain)
- Standardized reporting of side-effects and adverse events is needed

## Introduction

Neuromodulation, particularly noninvasive brain stimulation, including repetitive transcranial magnetic stimulation (rTMS), has emerged as a promising intervention for the treatment of substance use disorders (SUD) and their comorbidities (1). rTMS can modulate cortical excitability and network dynamics involved in craving, impulsivity, and maladaptive reward processing (2,3). Numerous randomized controlled trials have investigated the efficacy of TMS in tobacco, alcohol, stimulant, opioid, and cannabis use disorders. The clinical efficacy signals are encouraging. Recent comprehensive reviews and meta-analyses report that TMS can reduce craving and use in a variety of substance use disorders (4–6).

rTMS is generally considered safe when participants are properly screened and rTMS is applied within approved therapeutic protocols in many clinical populations (7). Studies investigating its use in psychiatric disorders such as depression, obsessive–compulsive disorder, schizophrenia, and post traumatic stress disorder have consistently shown it to be safe and well-tolerated (8–14). Seizure provocation has historically been considered as the most serious risk associated with rTMS; however, its incidence is very low (0.71 per 1,000 patients) when proper screening and dosing protocols are followed (15). Common, non-serious adverse effects include headache, scalp discomfort, and transient hearing changes (7,16–18). However, caution should be taken when applying any intervention to any sample.

Some consideration for safety is needed in SUD samples with and without co-occurring psychiatric conditions (15, 16) even though rTMS is generally well tolerated in SUD populations (21–23). However, there is a lack of systematic evaluations of the details, types, and incidence of adverse events (AEs) in published trials. Understanding the safety profile or rTMS in SUD samples could help such individuals who have been historically subject to stigma and barriers to access to evidence-based treatments (24). A list of recommendations and contraindications in a recently published guideline for rTMS in people with SUD has increased this concern (20), although this concern is not universal (25). To thoroughly evaluate the empirical evidence, we conducted a systematic review that aims to consolidate existing evidence on rTMS-related side effects and AEs in published trials in SUD populations, with attention to factors such as substance type, treatment status, stimulation target, and dose. We seek to empirically support the risk profile of applying rTMS to SUD samples which will better inform future trial designs, clinical decision-making, and the responsible integration of neuromodulation into evidence-based addiction treatment strategies.

## Methods

### Search Strategy

A systematic search of PubMed was conducted to identify studies published up to January 1, 2025, that evaluated rTMS as an intervention for SUDs. This review was conducted in accordance with the latest Preferred Reporting Items for Systematic Reviews and Meta-Analyses (PRISMA) guidelines (26). The search terms are presented in Table S1 of the supplement.

The initial search yielded 582 articles on rTMS. Two independent investigators (AS, SS) screened and extracted data, with a third reviewer (HE) resolving any discrepancies. Titles and abstracts were first reviewed to exclude book chapters, commentaries, author corrections, editorials, review articles, studies involving non-human subjects, studies published in languages other than English, and studies focusing on disorders other than SUDs. Duplicate studies were also excluded.

After this initial screening, 213 articles progressed to full-text review. Full-text screening identified 141 eligible studies. Exclusion criteria at this stage included studies involving only healthy participants, case reports or case series, study protocols, and electric field modeling studies. Studies using single- or paired-pulse TMS were also excluded. With these criteria, we reviewed published reports of rTMS in a variety of SUD samples and directly compared AEs in active vs sham groups; thus, this assessment is perhaps the strongest measurement of the safety and tolerability of rTMS in SUD samples. The PRISMA flowchart detailing the inclusion and exclusion process is presented in Figure S1 in the supplementary materials.

### Data Extraction

From each included study, the following information was extracted: type of substance used, number of participants in active arms, participants’ treatment status, number of rTMS sessions, primary stimulation site (coil placement), and the detailed rTMS parameters, including stimulation site, stimulation type, neuronavigation system, coil type, frequency, intensity, and total number of pulses. Participants’ treatment status at the time of intervention was categorized into four stages: Active use (not yet sought standard treatment), pre-treatment (treatment-seeking but not yet started standard intervention), early recovery (within the first month of standard treatment, primarily detoxification and stabilization), and sustained recovery (more than one month since initial recovery).

### Adverse Event Reporting

Adverse effects were extracted and categorized according to the level of reporting detail. Studies specifying the frequency, type, or severity of AEs were classified as having detailed AE reporting, while those providing only general information were classified as summary AE reporting. Studies explicitly reporting no adverse effects were labeled as “No AEs,” and those that did not report any information were labeled as “Not reported.” For safety outcomes, adverse effects were further classified as no AEs, mild AEs (transient effects), moderate AEs (did not require treatment but were reported as moderate by participants), severe AEs (rated as severe and sometimes resulting in discontinuation of intervention), or unspecified (no safety information was provided). For studies lacking AE data, authors were contacted via email up to three times, and information was obtained and incorporated into the dataset.

## Results

A total of 141 TMS studies involving 4,299 participants in active treatment arms met the inclusion criteria (see PRISMA flowchart in Figure S1 of the supplement). Studies were categorized by both substance type and treatment status (Figure 1). The largest proportion of studies enrolled pre-treatment, treatment-seeking individuals (61 studies, 2,331 participants). This was followed by active users (non-treatment-seeking) (37 studies, 888 participants), those in sustained recovery (25 studies, 627 participants), and participants in early recovery or detoxification (18 studies, 449 participants).

**Figure 1.**
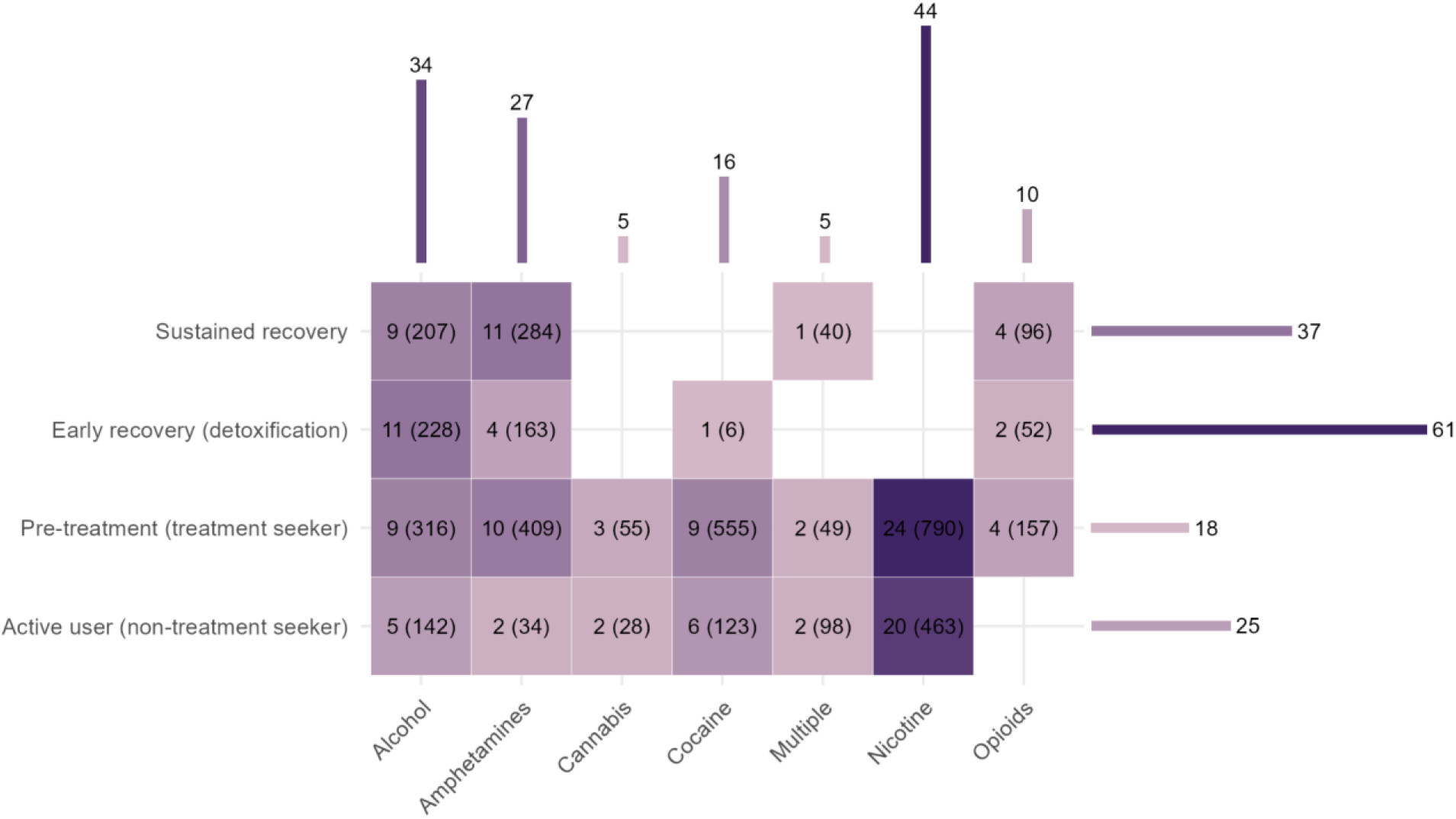
Variations in Substance Type and Treatment Status in TMS Studies for Substance Use Disorders. This figure presents the number of transcranial magnetic stimulation (TMS) studies on substance use disorders published through the end of 2024 (total studies = 141; total participants in active arms = 4,299), categorized by substance type and treatment status. Each title indicates the number of studies, with the total number of participants in the active arm shown in parentheses. The heatmap illustrates the distribution of studies across different substances and treatment statuses, with darker shades representing higher concentrations of published studies.

The included studies were conducted across six major substance groups. Nicotine (44 studies, 1,253 participants), alcohol (34 studies, 893 participants), and amphetamines (27 studies, 890 participants) were the most frequently investigated. Fewer studies targeted cocaine (16 studies, 684 participants), opioids (10 studies, 305 participants), or cannabis (5 studies, 83 participants). polysubstance use, defined as studies reporting outcomes for participants with more than one type of substance use, was examined in 5 studies (147 participants).

Alcohol and amphetamine studies were distributed across all treatment stages, though most were concentrated in pre-treatment and sustained recovery cohorts. Nicotine studies were predominantly conducted in pre-treatment (24 studies, 790 participants) and active-user samples (20 studies, 463 participants), reflecting the clinical need to intervene in nicotine use disorder before formal treatment engagement. Cocaine use disorder studies focused mainly on pre-treatment (9 studies, 555 participants), while opioid (4 studies, 157 participants) and cannabis research (3 studies, 55 participants) remained comparatively limited. The imbalance across treatment stages indicates that although TMS has been most frequently studied in treatment-seeking populations, opportunities remain to extend investigations to early recovery and sustained recovery populations.

A total of 141 studies were synthesized across substance type, treatment status, stimulation targets, dosing parameters, adverse event reporting, and safety outcomes (Figure 2). The dorsolateral prefrontal cortex (DLPFC) was the predominant stimulation target: 99 study arms targeted the left DLPFC with high-frequency stimulation (≥ 5 Hz); 10 study arms targeted the left DLPFC with low-frequency stimulation (≤ 1 Hz); 34 study arms targeted the right DLPFC with high-frequency stimulation; and 6 study arms targeted the right DLPFC with low-frequency stimulation. Less commonly, stimulation was directed at the frontopolar cortex (FPC; n=10), inferior frontal gyrus (n=3), insula (n=2), or superior frontal gyrus (n=1). Thirteen studies reported stimulation of more than one brain region. Dosing intensity varied considerably across studies. The most common protocol was 100% of the resting motor threshold (RMT) (n=59), followed by 110% RMT (n=35), 120% RMT (n=14), 80% RMT (n=16), and 90% RMT (n=11). Only one study used 70% RMT. Three studies applied more than one intensity, and two did not report any dosing information. Regarding the type of TMS coil used, the figure-of-8 coil was the most frequently reported (n=99), followed by H-coils (n=16), round coils (n=7), and double cone coils (n=1). The remaining studies did not specify the coil type used.

**Figure 2.**
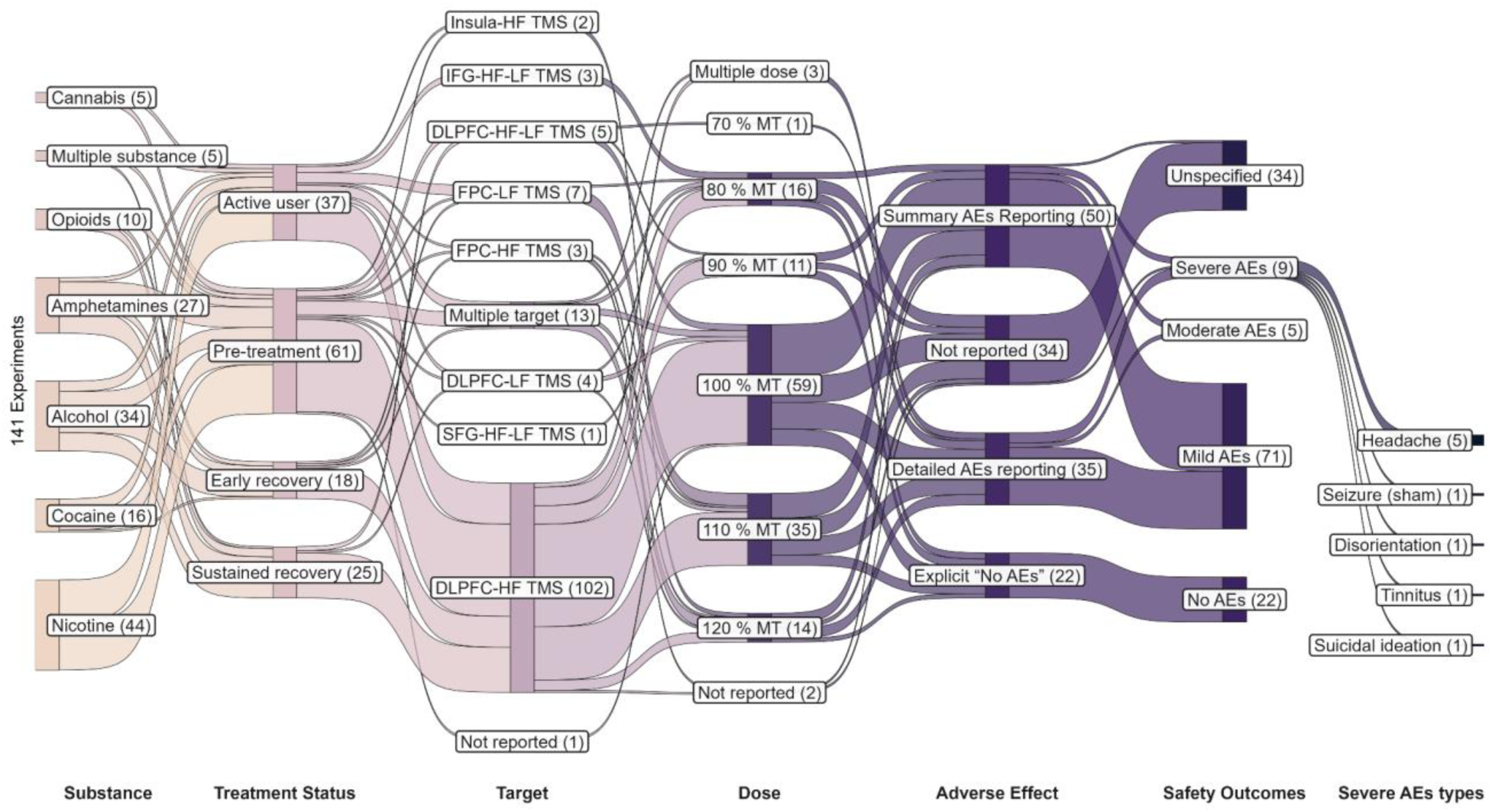
Substance type, treatment status, target areas, dose, adverse effect reporting, and outcomes in TMS studies for substance use disorders up to the end of 2024. This Sankey diagram illustrates the distribution of 141 TMS studies, including 4,299 participants in the active arm, categorized by substance type, treatment status, targeted brain regions, dose, adverse effect reporting, and outcomes. In the substance layer, Multiple refers to studies that included participants with more than one type of substance use. In the target area layer, Multiple refers to studies that employed stimulation at more than one site. Dose refers to TMS intensity, expressed as a percentage of the resting motor threshold; Multiple in the dose layer indicates studies that applied more than one stimulation dose. In the adverse effect reporting layer, summary AE reporting refers to studies that provided only general information on side effects without details on frequency, type, or severity (e.g., reporting only the type of side effect or a total count without group breakdown). Detailed AE reporting refers to studies that specify the frequency, type, and/or severity of adverse effects with comprehensive descriptions. Abbreviations: TMS – transcranial magnetic stimulation; DLPFC – dorsolateral prefrontal cortex; FPC – frontopolar cortex; SFG – superior frontal gyrus; IFG – inferior frontal gyrus; HF – high frequency; LF – low frequency; MT – motor threshold; AEs – adverse effects.

A majority of the studies either only offered a summary of AEs information (n=50, 35.46%) or did not report AEs (n=35, 24.82). In contrast, 34 studies (24.11%) provided detailed AE descriptions, and 22 (15.60%) explicitly stated that no adverse events occurred. Regarding safety outcomes, twenty-two studies (15.60%) described the interventions as safe, with no adverse effects. Most studies reported mild adverse events (n=69, 48.94%; e.g., mild headache) and only a few studies reported at least one moderate (n=6, 4.26%; e.g., moderate headache) or severe (n=9, 6.38%, e.g., tinnitus) AEs. Thirty-five studies did not report their safety outcomes, leaving them unspecified. Moderate and severe AEs were very uncommon but due to insufficient detail, the number of events is difficult to quantify. However, no rTMS-related seizures (the most severe AE) were reported in these 141 studies with 4,299 participants suggesting rTMS is safe in SUD samples with appropriate screening procedures.

Severe cases included the following: one seizure occurred in the DLPFC-HF rTMS sham group in a patient with alcohol use disorder who was within six days of discontinuing lorazepam, which had been administered during detoxification. This event led to study dropout (27). One study reported two severe adverse events among patients with alcohol use disorder receiving DLPFC iTBS. One event occurred in the sham group and involved homicidal ideation, while the other was in the active group and involved an episode of alcohol intoxication accompanied by suicidal ideation in a veteran with comorbid alcohol use disorder (28). Another case involved a nicotine-dependent patient who received deep TMS and developed tinnitus, which led to study discontinuation but later resolved (29). Additionally, one patient enrolled in a tobacco treatment study discontinued participation due to a confusional state, disorientation, and difficulty following commands. This episode was identified during the pre-session assessment at the third DLPFC-HF TMS visit and was determined to be unrelated to the TMS treatment (30). Five other studies reported cases of severe headache (88, 89, 90, 104, 110), and in four of these (31–34), the side effect led to dropout. Besides the reported side effects, most of which resulted in dropout, three additional studies reported participant dropout due to adverse effects. Headache was the most frequently cited reason (n=2) (35,36). Additionally, one study reported scalp pain in a participant from the active arm, leading to dropout (27). Only 9 studies reported individuals discontinuing due to rTMS discomfort suggesting rTMS is well tolerated in SUD samples. For a detailed summary of adverse events and study-specific information, see Table S2 of the supplement.

## Discussion

This systematic review provides an up-to-date summary of safety and AEs reported in the rapidly growing field of rTMS for the treatment of SUDs. We characterized the rate of reported AEs and reported SAEs in 141 clinical trials and 4,299 unique participants, providing a convincing case that rTMS is safe and well-tolerated. This evidence supports future applications of rTMS as a stand-alone or an adjuvant intervention SUDs.

Although we found that 75.18% of all studies reported information on safety outcomes, the field should require such information from all published reports. Thus, there should be a standard for reporting side effects and tolerability to guide clinicians, researchers, and policymakers. Importantly, in those that did report on safety outcomes, no rTMS-induced seizures were reported and most studies found no AEs (20.75%) or only mild and transient AEs (65.09%). Therefore, moderate AEs were rare, severe AEs were even more rare. These findings are in line with the consensus that rTMS is a relatively safe and tolerable procedure (25). In other conditions for which rTMS has a longer and broader history of use, such major depressive disorder, serious AEs are rare, and tolerability is high (37).

Ongoing accumulation and evaluation of safety data in patients with SUDs receiving rTMS is essential for providing up-to-date guidance for clinicians and researchers in the field. Current rTMS safety guidelines specifically identify the use of alcohol and substances as a risk factor for seizures (7). Seizures are generally thought to be the most serious and life-threatening AE from rTMS, and previous reports have suggested that it occurs at very low rates in all samples (33) and our review of 4,299 participants without a seizure suggests rTMS in SUD samples have a similar (or even lower) risk profile. Our review showed that no rTMS-induced seizures were reported. Overall, SUD samples properly screened for seizure risk factors (e.g., acute intoxication, alcohol withdrawal, use of benzodiazepines) (7,39) do not have elevated risk for rTMS-induced seizures, supporting a recent consensus (25). In this review, there is no supporting evidence to the thought that recent substance cessation in regular users increased rTMS risk as we reviewed 18 studies with 449 participants during the detoxification or early recovery period without a single seizure reported. Although this evidence does not rule out the chance of a seizure in such situations, the empirical evidence reviewed here suggest no elevated risk to individuals in these situations.

With respect to tolerability and the most common AEs, headaches were the most reported, which is in line with the literature on rTMS for all neuropsychiatric disorders (40). The incidence of headaches in this population with SUDs did not seem different than what is typically reported in other conditions (41). Similarly, the severity was typically mild to moderate and transient. We only found reports of 10 participants receiving active rTMS (not including those in sham conditions) across all studies in our review who dropped out as a direct result of an adverse event. These findings suggest that even in the presence of common AEs, rTMS is well tolerated in SUD samples.

Limitations of such a review reflect the current landscape of rTMS and SUD research. There is an underrepresentation of certain SUDs; more data is required for rTMS of cocaine, cannabis, and opioid use disorders. Furthermore, the included studies represent controlled clinical trials, typically from academic or research settings with few coming from clinical settings. Most of the studies did not report the exact number of participants experiencing each specific side effect, making it impossible to calculate reliable prevalence estimates for each AE. Standardization in reporting is needed to fully characterize the AE profile.

In conclusion, our review found that rTMS was very safe and tolerable for patients with SUDs, regardless of substance or phase of recovery. Our review is the first to summarize all randomized, controlled trials to date. Moving forward, the field should standardize reporting of rTMS side-effects to fully capture the risk profile of rTMS across SUD samples. More research is needed in specific samples (e.g., cocaine, cannabis, opioid use disorder) to bolster our conclusions about safety and tolerability across SUD samples. An interesting future direction will be to explore tolerability differences across specific rTMS parameters such as dose, timing of stimulation delivery, and simulation target to rapidly identify the optimal rTMS intervention for SUDs.

## Data Availability

All data produced in the present work are contained in the manuscript

## Acknowledgment

We gratefully acknowledge the authors who kindly responded to our email inquiries and provided additional details on side effects in their clinical trials, including Alessandra Del Felice, Amedeo Amedei, Biswa Ranjan Mishra, Brett Froeliger, Christine Sheffer, Francesca Filbey, Georgios Mikellides, Hang Su, Hui Zheng, Jochem Milan Jansen, K J Divinakumar, Min Zhao, Nicole Petersen, Ti-Fei Yuan, Tony George, Travis Baker, William V Lechner, and Yixuan Ku. Their contributions were invaluable in ensuring the accuracy and completeness of our review.

## Supplementary materials

**Figure S1.**
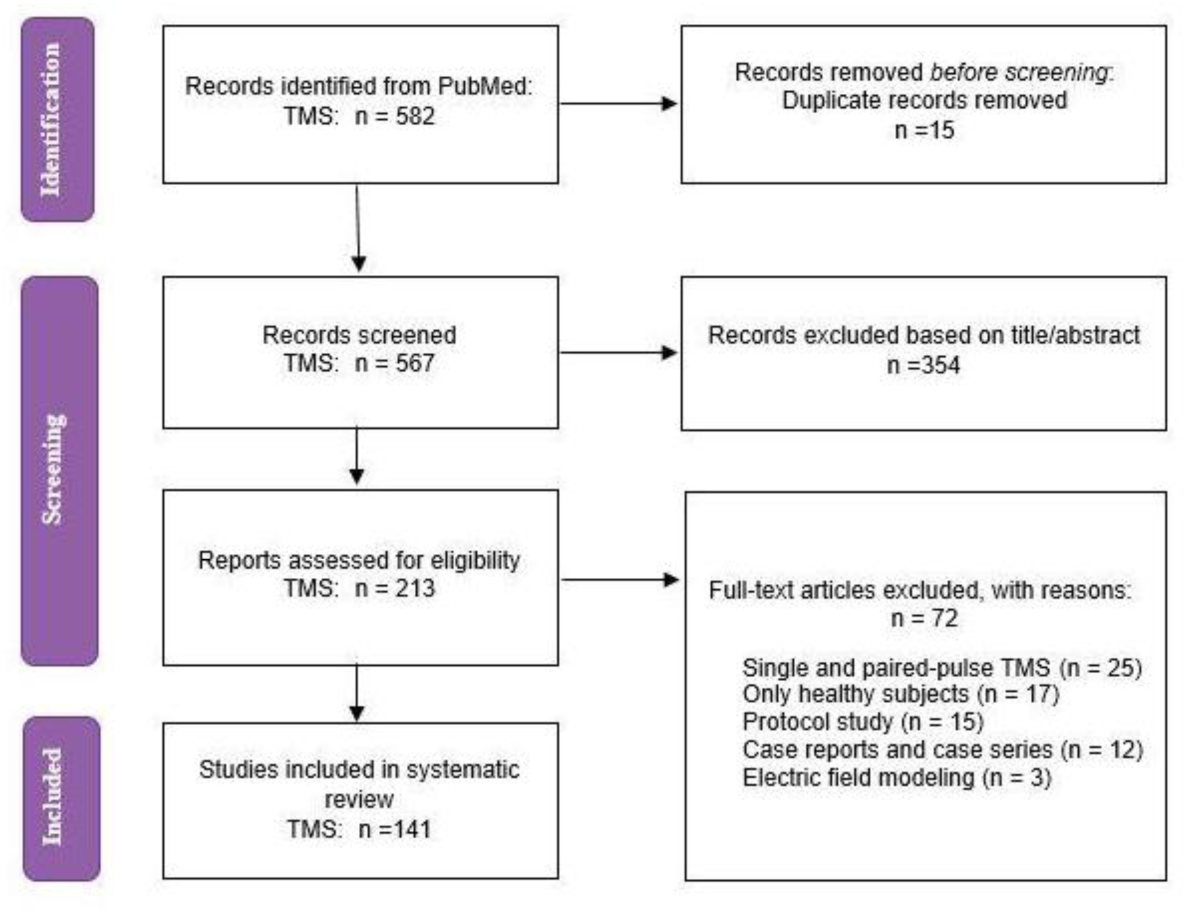
PRISMA Flowcharts for TMS Studies in Substance Use Disorders. This PRISMA flow diagram represents published trials of TMS for individuals with substance use disorders from inception up to 1 January 2025.

**Table S1.**
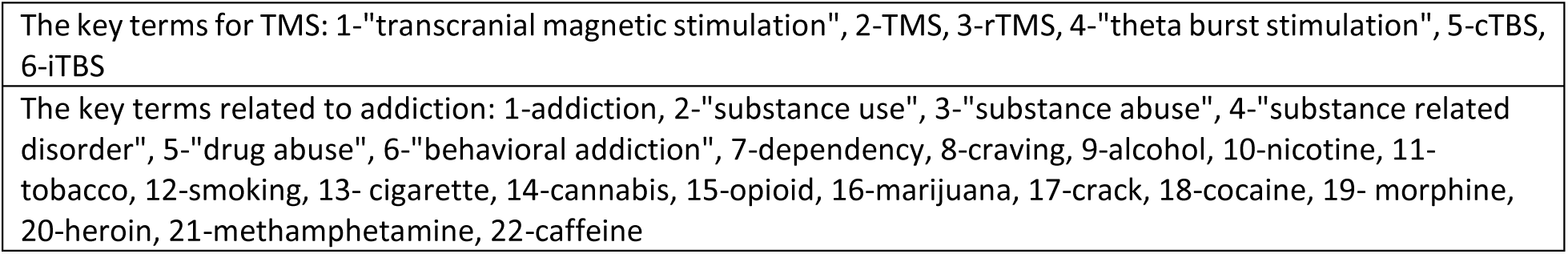
Key terms used to search in the PubMed database.

**Table S2.**
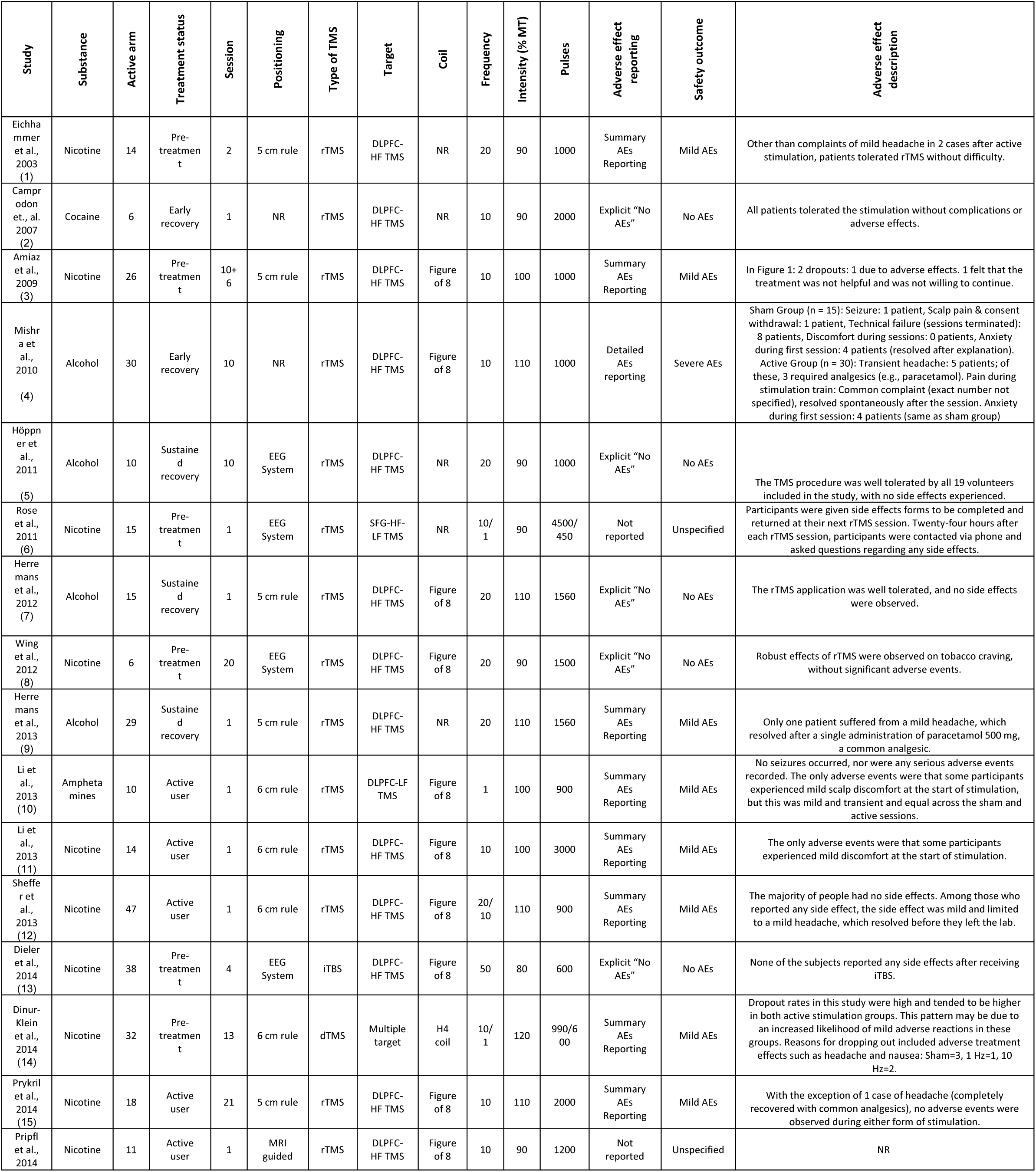
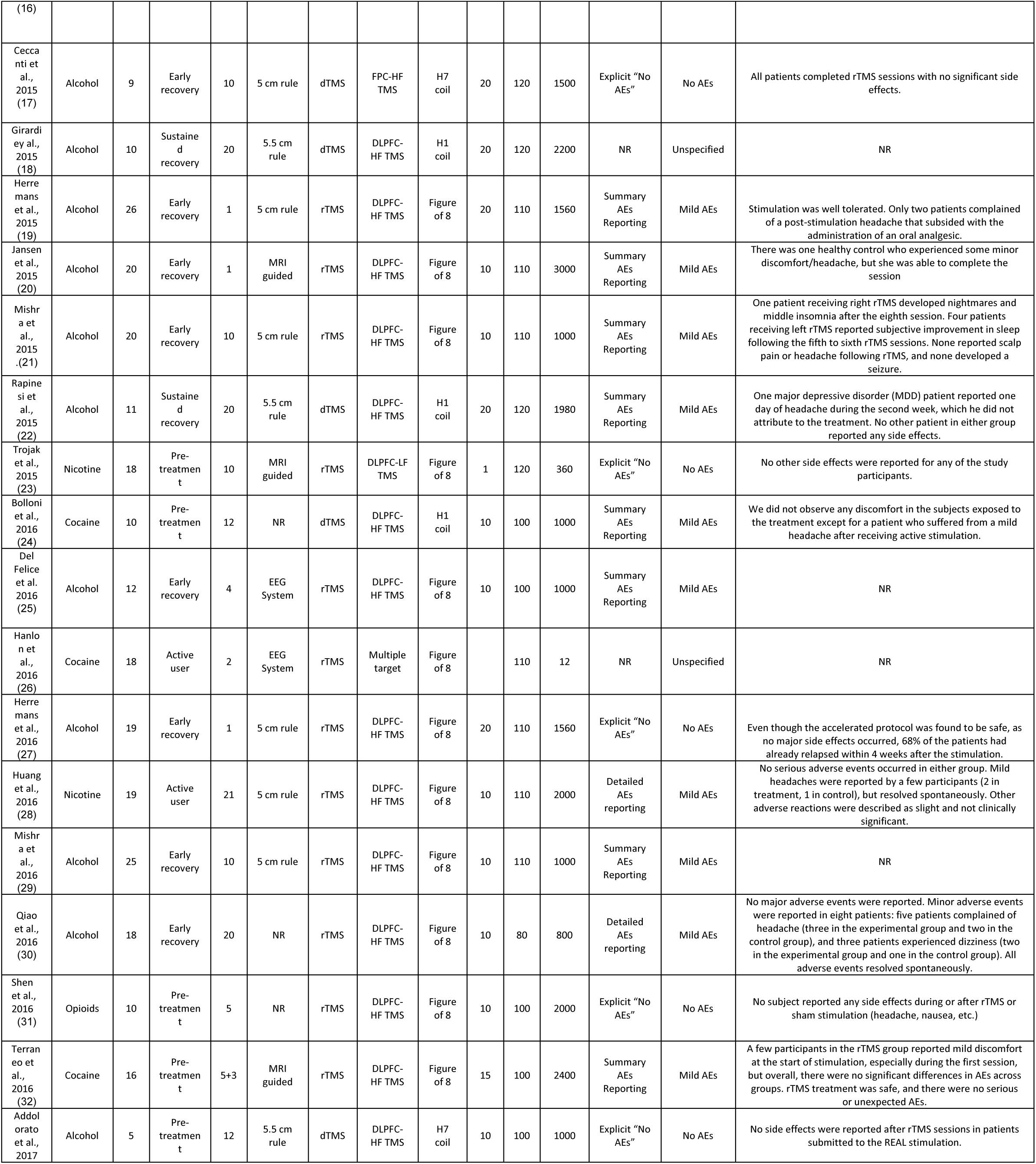
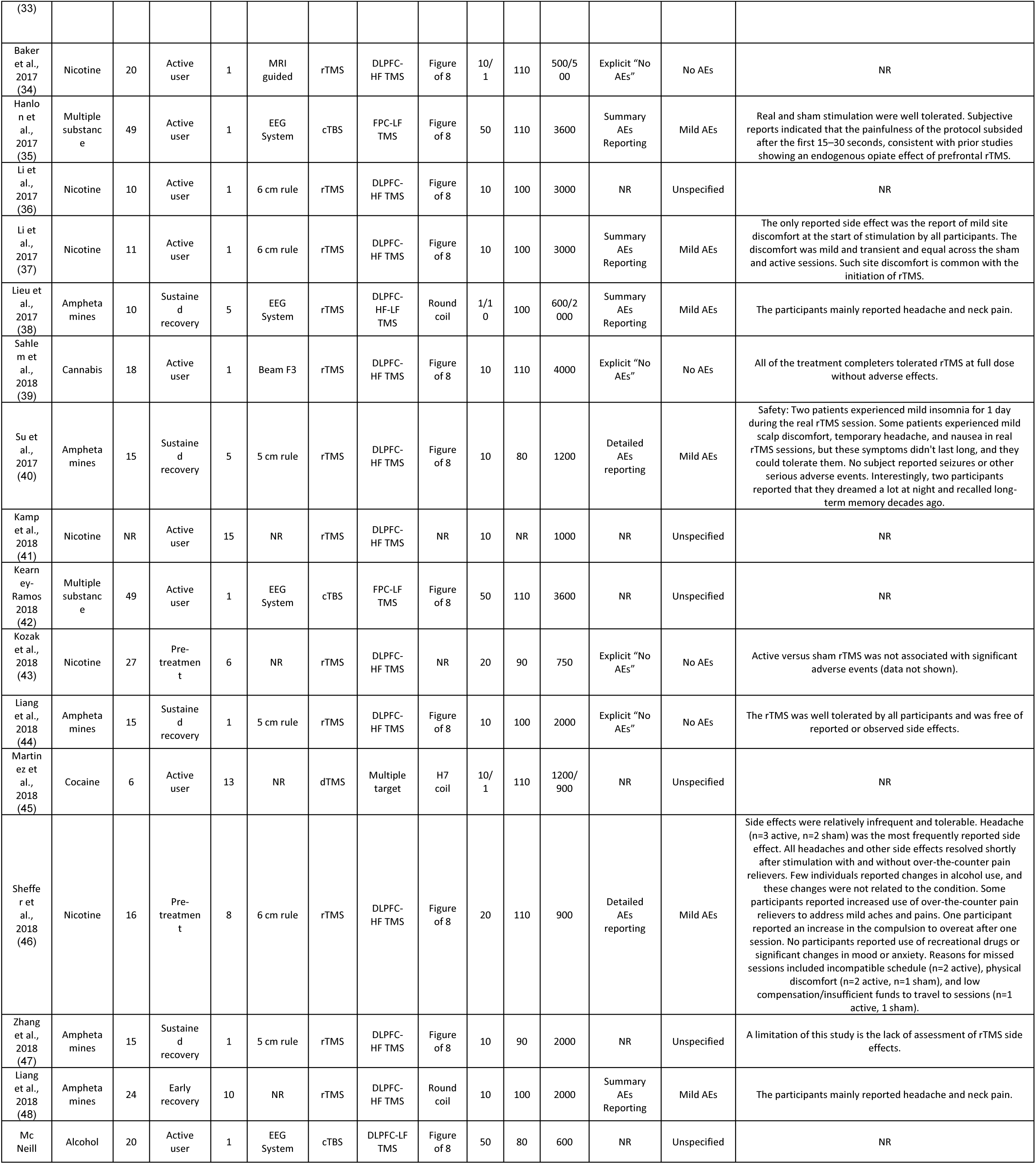
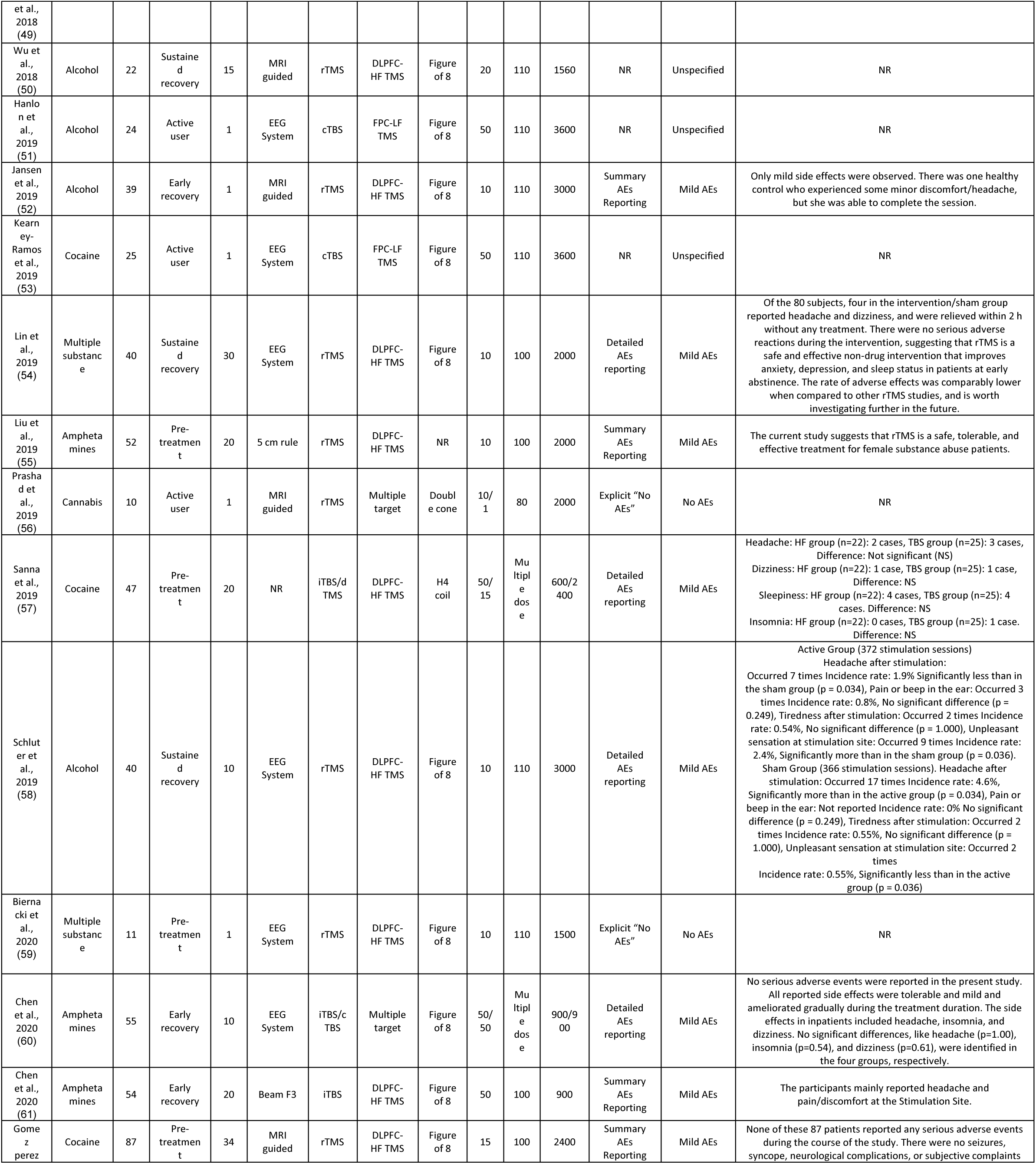
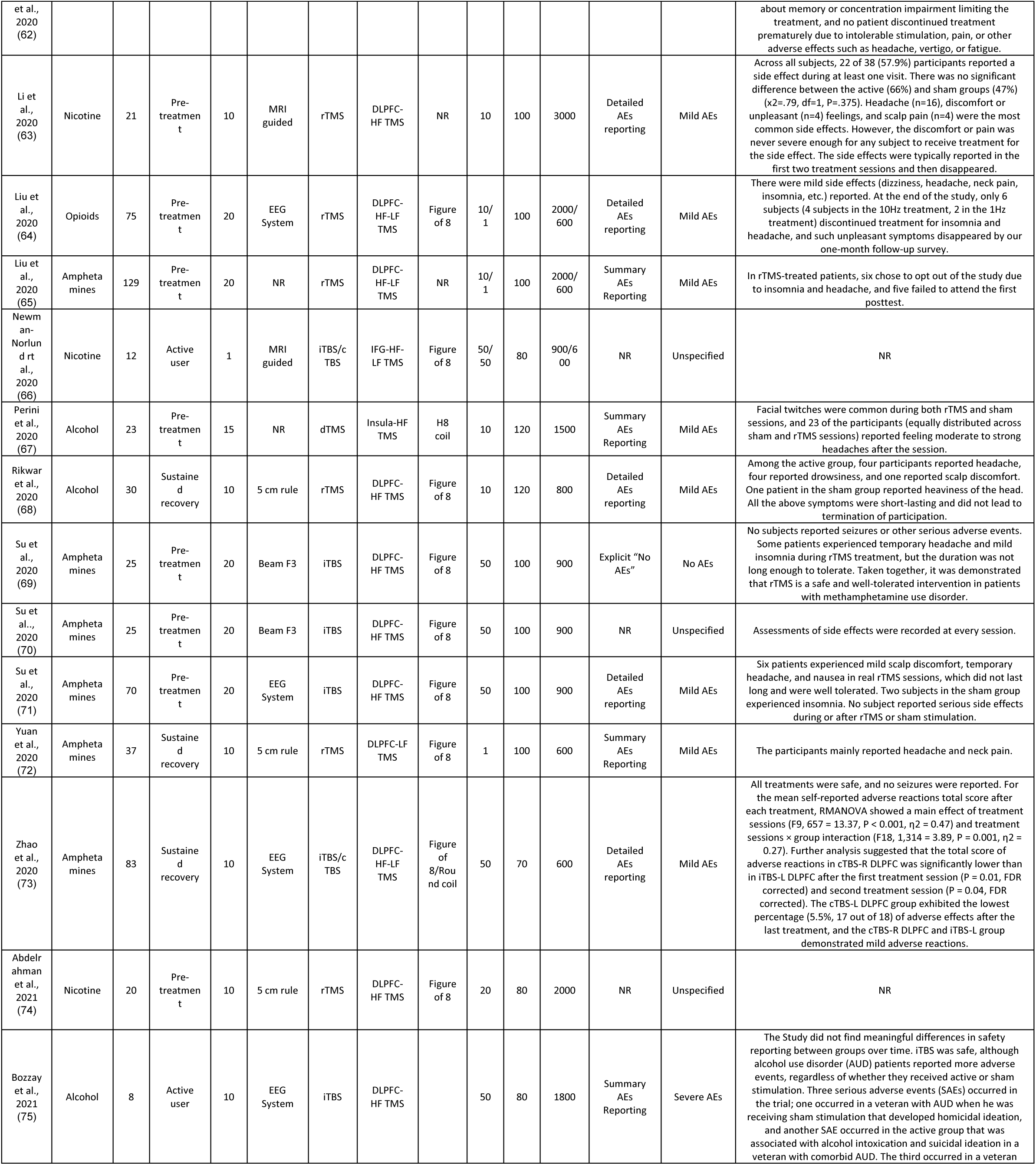
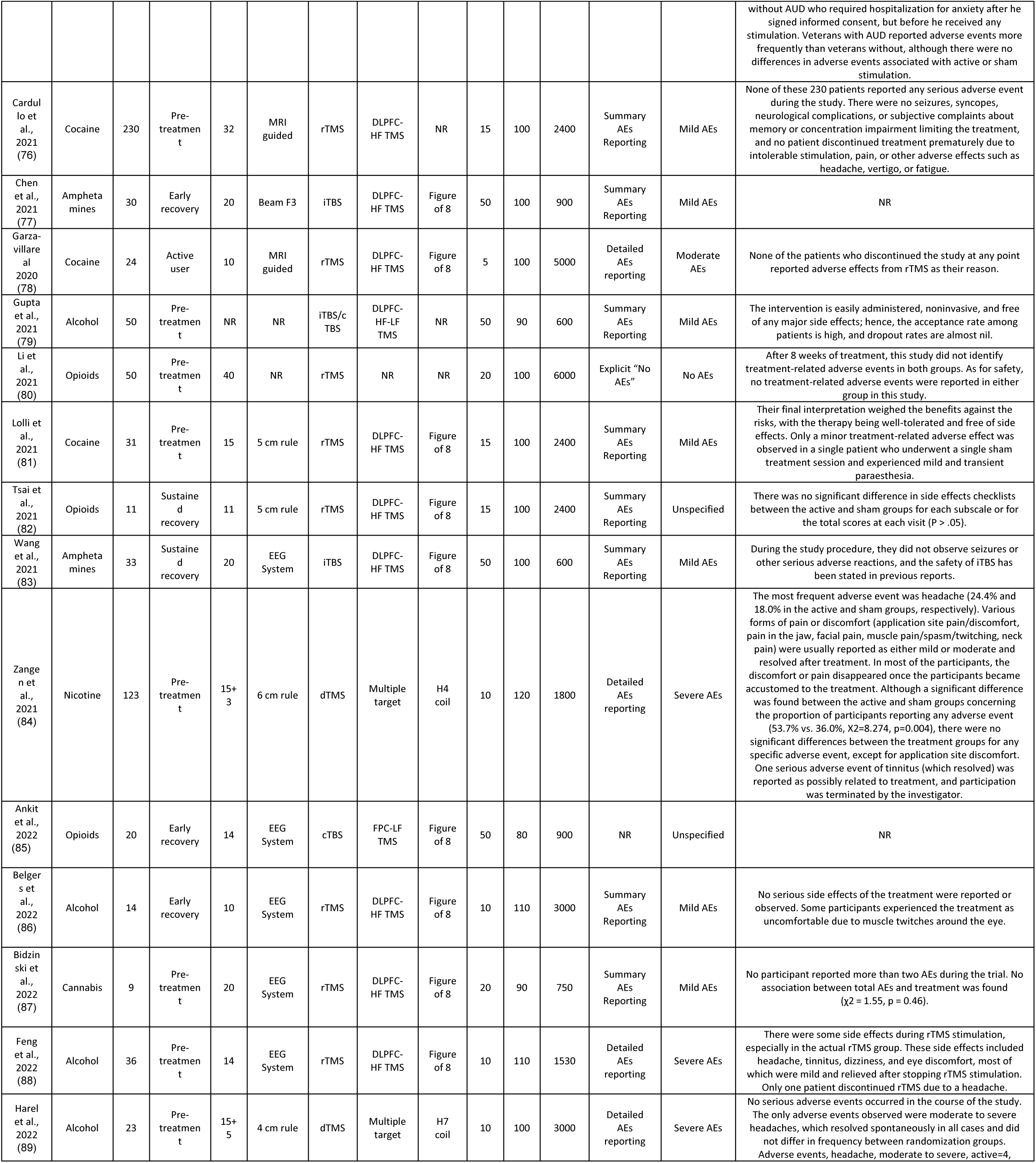
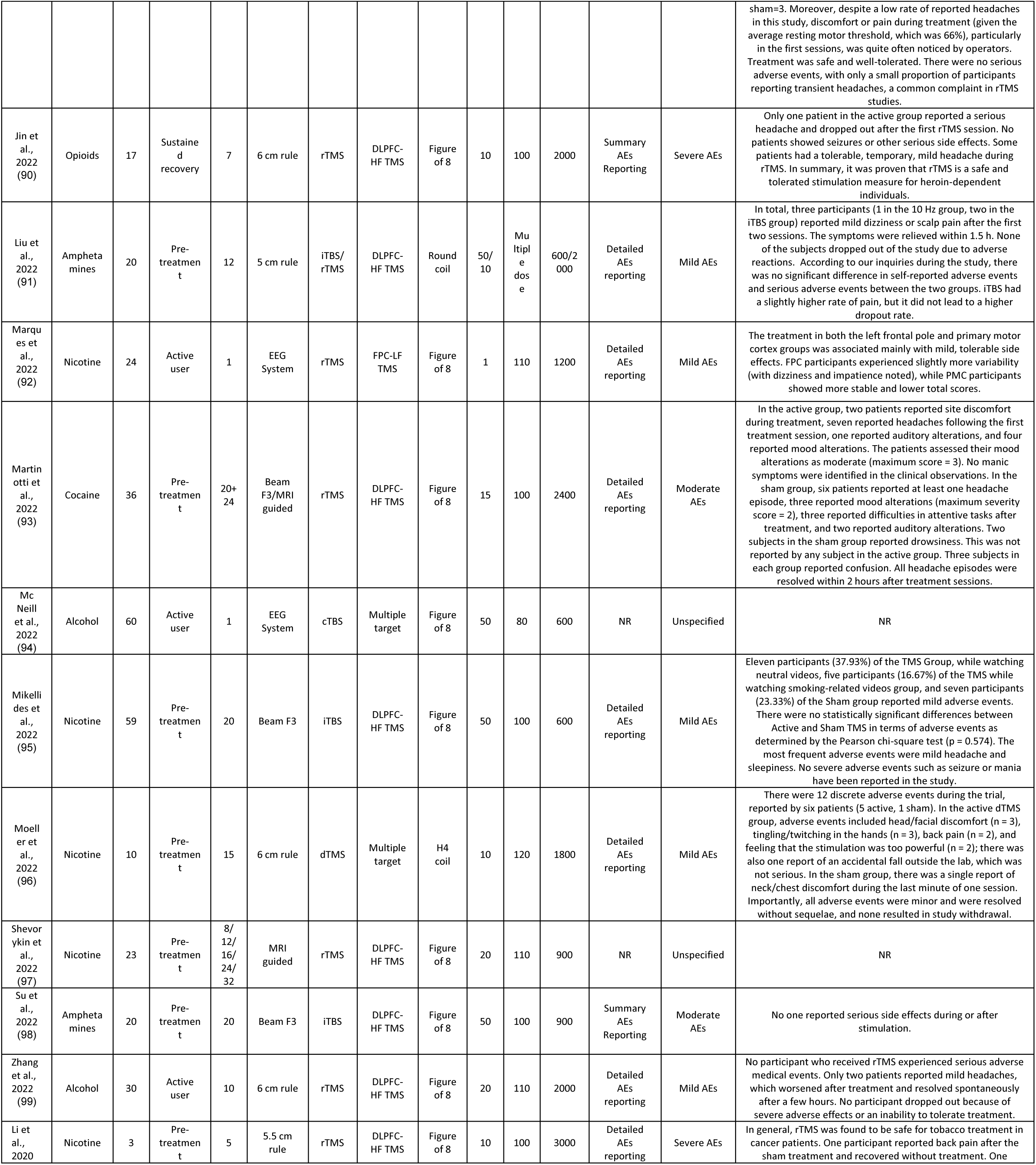
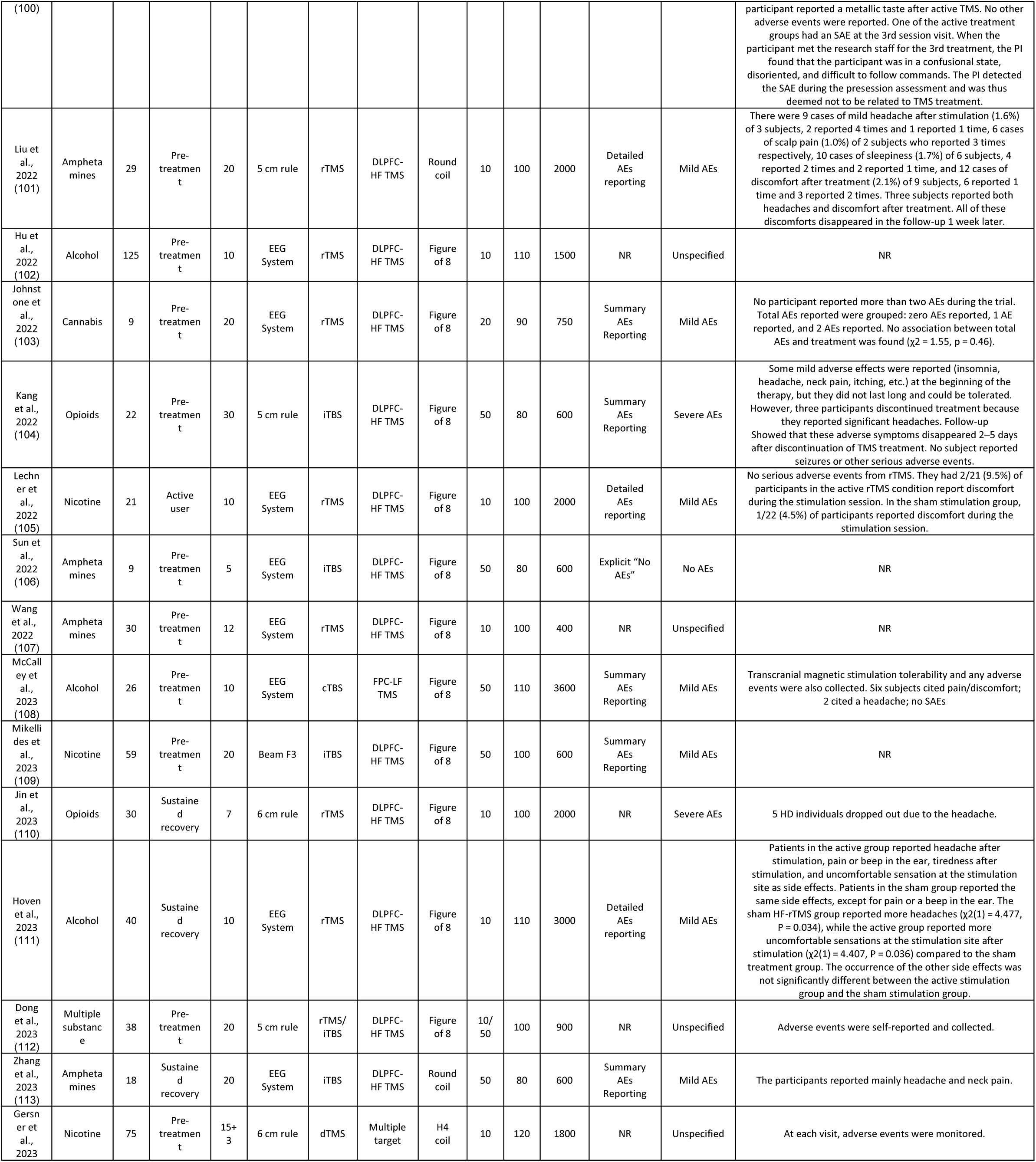
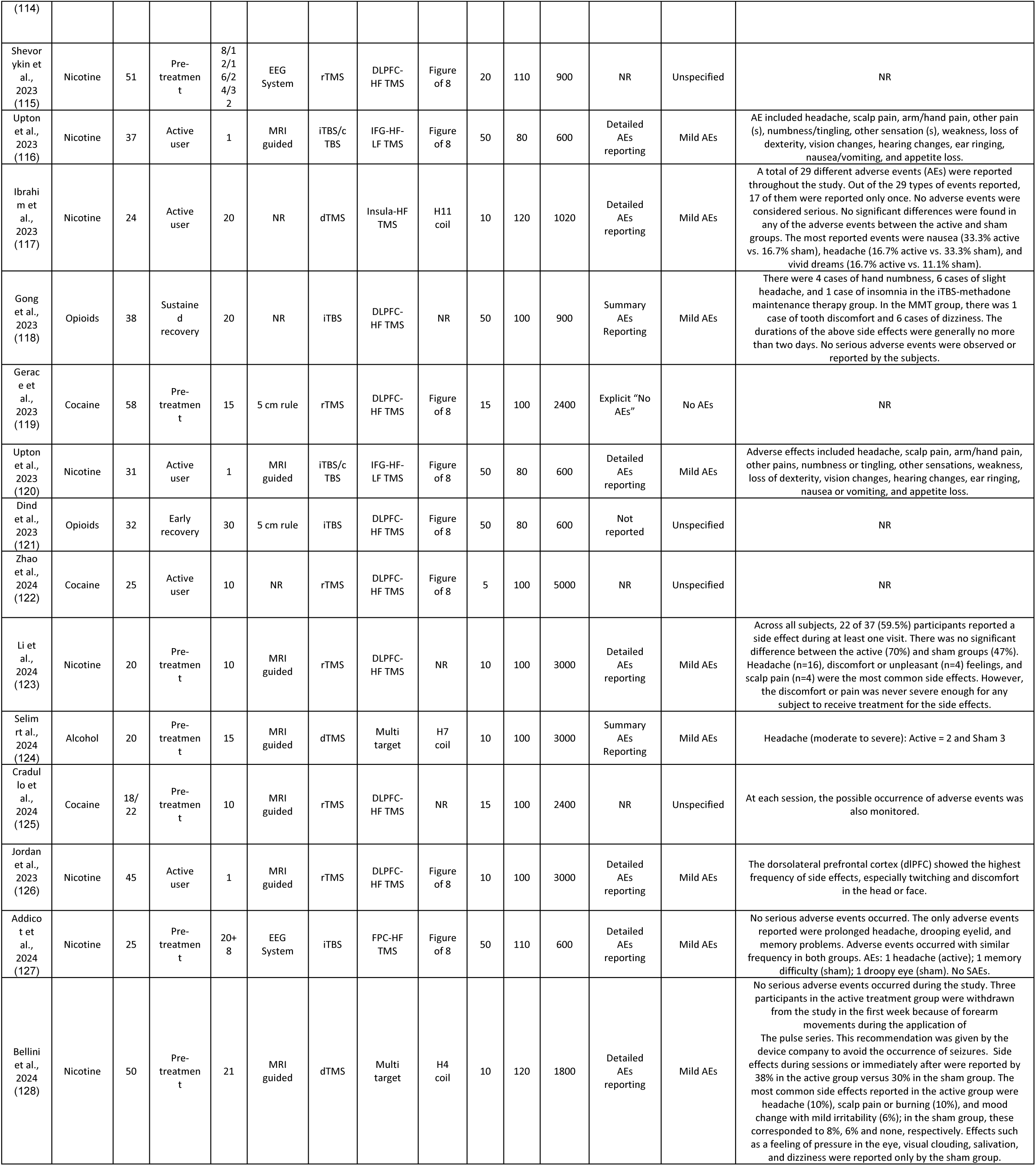
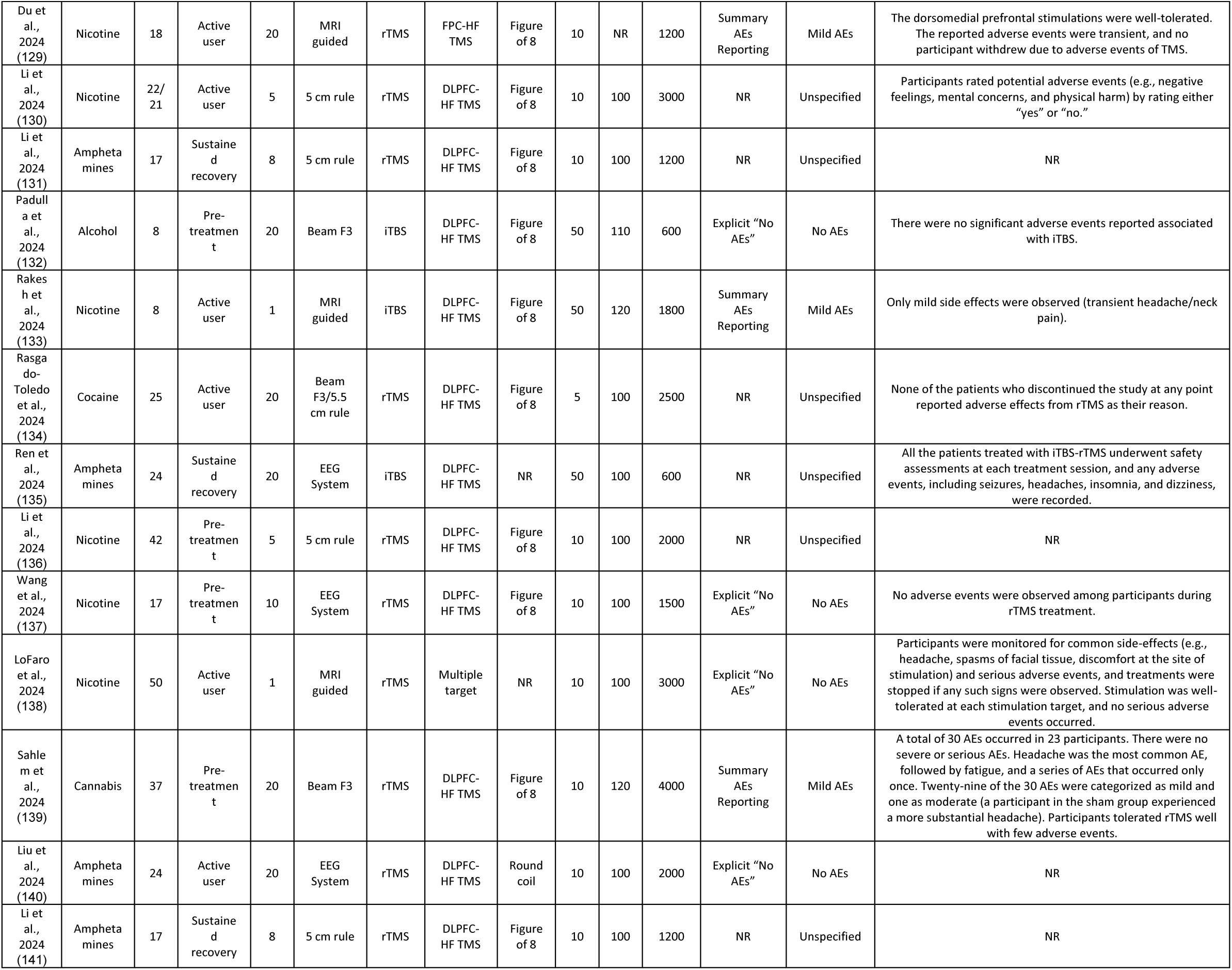
Summary of TMS protocol parameters and reported safety outcomes in studies on substance use disorders. This table presents key intervention characteristics, including stimulation parameters (TMS type, cortical target, positioning system, coil type, frequency, intensity, pulse count), experimental design elements (substance studied, sample size in active arm, treatment status, number of sessions), and safety reporting methods (adverse event monitoring, safety outcomes, and descriptions of specific adverse events). The structured format facilitates cross-study comparison of TMS safety profiles and supports protocol standardization efforts. **Abbreviations:** TMS, transcranial magnetic stimulation; HF TMS, high-frequency TMS (≥ 5 Hz); LF TMS, low-frequency TMS (≤ 1 Hz); rTMS, repetitive TMS; dTMS, deep TMS; iTBS, intermittent theta-burst stimulation; CTBS, continuous theta-burst stimulation; EEG, electroencephalography; MRI, magnetic resonance imaging; DLPFC, dorsolateral prefrontal cortex; FPC, frontopolar cortex; SFG, superior frontal gyrus; IFG, inferior frontal gyrus; AEs, adverse events; NR, not reported.

## References

1. Sage Reference - The Sage Handbook of Addiction Psychology - Neuromodulation (Brain Stimulation) Technologies for Treatment of Substance Use Disorders [Internet]. [cited 2025 Oct 2]. Available from: https://sk.sagepub.com/hnbk/edvol/the-sage-handbook-of-addiction-psychology/chpt/37-neuromodulation-brain-stimulation-technologies

2. Ekhtiari H, Tavakoli H, Addolorato G, Baeken C, Bonci A, Campanella S, et al. Transcranial electrical and magnetic stimulation (tES and TMS) for addiction medicine: A consensus paper on the present state of the science and the road ahead. Neurosci Biobehav Rev. 2019 Sept;104:118–40.

3. Soleimani G, Conelea CA, Kuplicki R, Opitz A, Lim KO, Paulus MP, et al. Targeting VMPFC-amygdala circuit with TMS in substance use disorder: A mechanistic framework. Addiction Biology. 2025 Jan;30(1):e70011.

4. 4. Soleimani G, Souki A, Honari S, Baker TE, Brunoni AR, Ebrahimi M, et al. Effectiveness of Noninvasive Brain Stimulation Protocols on Drug Craving and Consumption/Relapse in Substance Use Disorders: A Systematic Review and Meta-analysis of 208 Clinical Trials and 36 Protocols [Internet]. medRxiv; 2025 [cited 2025 Sept 22]. p. 2025.09.21.25335559. Available from: https://www.medrxiv.org/content/10.1101/2025.09.21.25335559v1

5. Mehta DD, Siddiqui S, Ward HB, Steele VR, Pearlson GD, George TP. Functional and structural effects of repetitive transcranial magnetic stimulation (rTMS) for the treatment of auditory verbal hallucinations in schizophrenia: A systematic review. Schizophr Res. 2024 May;267:86–98.

6. Michel A, Kang D, Balderston N, Bond D, Steele V. Repetitive Transcranial Magnetic Stimulation for Substance Use Disorders and Chronic Pain: a Review of the Evidence and Call for Increased Mechanistic Understanding. Current Addiction Reports. 2025 Feb 7;12.

7. Rossi S, Antal A, Bestmann S, Bikson M, Brewer C, Brockmöller J, et al. Safety and recommendations for TMS use in healthy subjects and patient populations, with updates on training, ethical and regulatory issues: Expert Guidelines. Clin Neurophysiol. 2021 Jan;132(1):269–306.

8. Wang WL, Wang SY, Hung HY, Chen MH, Juan CH, Li CT. Safety of transcranial magnetic stimulation in unipolar depression: A systematic review and meta-analysis of randomized-controlled trials. J Affect Disord. 2022 Mar 15;301:400–25.

9. Janicak PG, O’Reardon JP, Sampson SM, Husain MM, Lisanby SH, Rado JT, et al. Transcranial magnetic stimulation in the treatment of major depressive disorder: a comprehensive summary of safety experience from acute exposure, extended exposure, and during reintroduction treatment. J Clin Psychiatry. 2008 Feb;69(2):222–32.

10. Edinoff AN, Hegefeld TL, Petersen M, Patterson JC, Yossi C, Slizewski J, et al. Transcranial Magnetic Stimulation for Post-traumatic Stress Disorder. Front Psychiatry. 2022;13:701348.

11. Blyth SH, Cruz Bosch C, Raffoul JJ, Chesley J, Johnson B, Borodge D, et al. Safety of rTMS for Schizophrenia: A Systematic Review and Meta-analysis. Schizophr Bull. 2025 Mar 14;51(2):392–400.

12. Overvliet GM, Jansen RAC, van Balkom AJLM, van Campen DC, Oudega ML, van der Werf YD, et al. Adverse events of repetitive transcranial magnetic stimulation in older adults with depression, a systematic review of the literature. Int J Geriatr Psychiatry. 2021 Mar;36(3):383–92.

13. S B, Ni B, A H, K K, A W, D S, et al. Efficacy and safety of transcranial magnetic stimulation for treating major depressive disorder: An umbrella review and re-analysis of published meta-analyses of randomised controlled trials. Clinical psychology review [Internet]. 2023 Mar [cited 2025 Sept 20];100. Available from: https://pubmed.ncbi.nlm.nih.gov/36587461/

14. Fitzsimmons SMDD, van der Werf YD, van Campen AD, Arns M, Sack AT, Hoogendoorn AW, et al. Repetitive transcranial magnetic stimulation for obsessive-compulsive disorder: A systematic review and pairwise/network meta-analysis. J Affect Disord. 2022 Apr 1;302:302–12.

15. Taylor JJ, Newberger NG, Stern AP, Phillips A, Feifel D, Betensky RA, et al. Seizure risk with repetitive TMS: Survey results from over a half-million treatment sessions. Brain Stimul. 2021;14(4):965–73.

16. Kim WS, Paik NJ. Safety Review for Clinical Application of Repetitive Transcranial Magnetic Stimulation. Brain Neurorehabil. 2021 Mar;14(1):e6.

17. Taylor JJ, Newberger NG, Stern AP, Phillips A, Feifel D, Betensky RA, et al. Seizure risk with repetitive TMS: Survey results from over a half-million treatment sessions. Brain Stimulation: Basic, Translational, and Clinical Research in Neuromodulation. 2021 July 1;14(4):965–73.

18. Tseng PT, Zeng BY, Hsu CW, Liang CS, Carvalho AF, Brunoni AR, et al. The non-invasive brain or nerve stimulation treatment did not increase seizure frequency in patients with epilepsy: A network meta-analysis. Epilepsy Behav. 2025 Mar;164:110290.

19. Gorelick DA, Zangen A, George MS. Transcranial magnetic stimulation (TMS) in the treatment of substance addiction. Ann N Y Acad Sci. 2014 Oct;1327(1):79–93.

20. Tang VM, Aaronson S, Abdelghani M, Baeken C, Barbour T, Brunoni AR, et al. Assessment and Management of Concurrent Substance Use in Patients Receiving Repetitive Transcranial Magnetic Stimulation for Depressive, Obsessive-Compulsive, Psychotic, and Trauma-Related Disorders: A Delphi Consensus Study and Guideline. Am J Psychiatry. 2025 May 1;182(5):438–51.

21. Madeo G, Terraneo A, Cardullo S, Gómez Pérez LJ, Cellini N, Sarlo M, et al. Long-Term Outcome of Repetitive Transcranial Magnetic Stimulation in a Large Cohort of Patients With Cocaine-Use Disorder: An Observational Study. Front Psychiatry. 2020;11:158.

22. Torres-Castaño A, Rivero-Santana A, Perestelo-Pérez L, Duarte-Díaz A, Toledo-Chávarri A, Ramos-García V, et al. Transcranial Magnetic Stimulation for the Treatment of Cocaine Addiction: A Systematic Review. J Clin Med. 2021 Nov 28;10(23):5595.

23. Tang VM, Le Foll B, Daskalakis ZJ, Wang AL, Buckley L, Blumberger DM, et al. Repetitive transcranial magnetic stimulation for the treatment of suicidality in opioid use disorder: a pilot feasibility randomized controlled trial. Eur Psychiatry. 2025 Mar 5;68(1):e63.

24. Richter L, Vuolo L, Salmassi MS. Stigma and Addiction Treatment. In: Avery JD, Avery JJ, editors. The Stigma of Addiction: An Essential Guide [Internet]. Cham: Springer International Publishing; 2019 [cited 2025 Sept 27]. p. 93–130. Available from: 10.1007/978-3-030-02580-9_7

25. Steele VR, Addicott MA, Addolorato G, Baker T, Bonci A, Brady KT, et al. Toward Inclusive, Evidence-Based rTMS Care for Patients with Co-Occurring Substance Use Disorders. American Journal of Psychiatry. (in press); 2025.

26. Page MJ, Moher D, Bossuyt PM, Boutron I, Hoffmann TC, Mulrow CD, et al. PRISMA 2020 explanation and elaboration: updated guidance and exemplars for reporting systematic reviews. BMJ. 2021 Mar 29;n160.

27. Mishra BR, Nizamie SH, Das B, Praharaj SK. Efficacy of repetitive transcranial magnetic stimulation in alcohol dependence: a sham-controlled study. Addiction. 2010 Jan;105(1):49–55.

28. Bozzay ML, Brigido S, van ’t Wout-Frank M, Aiken E, Swift R, Philip NS. Intermittent Theta Burst Stimulation in Veterans with Mild Alcohol Use Disorder. J Affect Disord. 2021 Oct 1;293:314–9.

29. Zangen A, Moshe H, Martinez D, Barnea-Ygael N, Vapnik T, Bystritsky A, et al. Repetitive transcranial magnetic stimulation for smoking cessation: a pivotal multicenter double-blind randomized controlled trial. World Psychiatry. 2021 Oct;20(3):397–404.

30. Li X, Toll BA, Carpenter MJ, Nietert PJ, Dancy M, George MS. Repetitive Transcranial Magnetic Stimulation for Tobacco Treatment in Cancer Patients: A Preliminary Report of a One-Week Treatment. J Smok Cessat. 2022;2022:2617146.

31. Feng Z, Wu Q, Wu L, Zeng T, Yuan J, Wang X, et al. Effect of High-Frequency Repetitive Transcranial Magnetic Stimulation on Visual Selective Attention in Male Patients With Alcohol Use Disorder After the Acute Withdrawal. Front Psychiatry. 2022;13:869014.

32. Jin L, Yuan M, Zhang W, Su H, Wang F, Zhu J, et al. Repetitive transcranial magnetic stimulation modulates coupling among large-scale brain networks in heroin-dependent individuals: A randomized resting-state functional magnetic resonance imaging study. Addict Biol. 2022 Mar;27(2):e13121.

33. Kang T, Ding X, Zhao J, Li X, Xie R, Jiang H, et al. Influence of improved behavioral inhibition on decreased cue-induced craving in heroin use disorder: A preliminary intermittent theta burst stimulation study. J Psychiatr Res. 2022 Aug;152:375–83.

34. Jin L, Yuan M, Zhang W, Wang L, Chen J, Wang F, et al. Default mode network mechanisms of repeated transcranial magnetic stimulation in heroin addiction. Brain Imaging Behav. 2023 Feb;17(1):54–65.

35. Amiaz R, Levy D, Vainiger D, Grunhaus L, Zangen A. Repeated high-frequency transcranial magnetic stimulation over the dorsolateral prefrontal cortex reduces cigarette craving and consumption. Addiction. 2009 Apr;104(4):653–60.

36. Dinur-Klein L, Dannon P, Hadar A, Rosenberg O, Roth Y, Kotler M, et al. Smoking cessation induced by deep repetitive transcranial magnetic stimulation of the prefrontal and insular cortices: a prospective, randomized controlled trial. Biol Psychiatry. 2014 Nov 1;76(9):742–9.

37. Trapp NT, Purgianto A, Taylor JJ, Singh MK, Oberman LM, Mickey BJ, et al. Consensus review and considerations on TMS to treat depression: A comprehensive update endorsed by the National Network of Depression Centers, the Clinical TMS Society, and the International Federation of Clinical Neurophysiology. Clin Neurophysiol. 2025 Feb;170:206–33.

38. Stultz DJ, Osburn S, Burns T, Pawlowska-Wajswol S, Walton R. Transcranial Magnetic Stimulation (TMS) Safety with Respect to Seizures: A Literature Review. Neuropsychiatr Dis Treat. 2020;16:2989–3000.

39. Tang VM, Ibrahim C, Rodak T, Goud R, Blumberger DM, Voineskos D, et al. Managing substance use in patients receiving therapeutic repetitive transcranial magnetic stimulation: A scoping review. Neurosci Biobehav Rev. 2023 Dec;155:105477.

40. Zis P, Shafique F, Hadjivassiliou M, Blackburn D, Venneri A, Iliodromiti S, et al. Safety, Tolerability, and Nocebo Phenomena During Transcranial Magnetic Stimulation: A Systematic Review and Meta-Analysis of Placebo-Controlled Clinical Trials. Neuromodulation. 2020 Apr;23(3):291–300.

41. Humaira A, Gao S, Gregory E, Ridgway L, Blumberger DM, Downar J, et al. A patient-oriented analysis of pain side effect: A step to improve the patient’s experience during rTMS? Brain Stimul. 2021;14(5):1147–53.

## References

1. Eichhammer P, Johann M, Kharraz A, Binder H, Pittrow D, Wodarz N, et al. High-frequency repetitive transcranial magnetic stimulation decreases cigarette smoking. J Clin Psychiatry. 2003 Aug;64(8):951–3.

2. Camprodon JA, Martínez-Raga J, Alonso-Alonso M, Shih MC, Pascual-Leone A. One session of high frequency repetitive transcranial magnetic stimulation (rTMS) to the right prefrontal cortex transiently reduces cocaine craving. Drug Alcohol Depend. 2007 Jan 5;86(1):91–4.

3. Amiaz R, Levy D, Vainiger D, Grunhaus L, Zangen A. Repeated high-frequency transcranial magnetic stimulation over the dorsolateral prefrontal cortex reduces cigarette craving and consumption. Addiction. 2009 Apr;104(4):653–60.

4. Mishra BR, Nizamie SH, Das B, Praharaj SK. Efficacy of repetitive transcranial magnetic stimulation in alcohol dependence: a sham-controlled study. Addiction. 2010 Jan;105(1):49–55.

5. Höppner J, Broese T, Wendler L, Berger C, Thome J. Repetitive transcranial magnetic stimulation (rTMS) for treatment of alcohol dependence. World J Biol Psychiatry. 2011 Sept;12 Suppl 1:57–62.

6. Rose JE, McClernon FJ, Froeliger B, Behm FM, Preud’homme X, Krystal AD. Repetitive transcranial magnetic stimulation of the superior frontal gyrus modulates craving for cigarettes. Biol Psychiatry. 2011 Oct 15;70(8):794–9.

7. Herremans SC, Baeken C, Vanderbruggen N, Vanderhasselt MA, Zeeuws D, Santermans L, et al. No influence of one right-sided prefrontal HF-rTMS session on alcohol craving in recently detoxified alcohol-dependent patients: results of a naturalistic study. Drug Alcohol Depend. 2012 Jan 1;120(1–3):209–13.

8. Wing VC, Bacher I, Wu BS, Daskalakis ZJ, George TP. High frequency repetitive transcranial magnetic stimulation reduces tobacco craving in schizophrenia. Schizophr Res. 2012 Aug;139(1–3):264–6.

9. Herremans SC, Vanderhasselt MA, De Raedt R, Baeken C. Reduced intra-individual reaction time variability during a Go-NoGo task in detoxified alcohol-dependent patients after one right-sided dorsolateral prefrontal HF-rTMS session. Alcohol Alcohol. 2013;48(5):552–7.

10. Li X, Malcolm RJ, Huebner K, Hanlon CA, Taylor JJ, Brady KT, et al. Low frequency repetitive transcranial magnetic stimulation of the left dorsolateral prefrontal cortex transiently increases cue-induced craving for methamphetamine: a preliminary study. Drug Alcohol Depend. 2013 Dec 1;133(2):641–6.

11. Li X, Hartwell KJ, Owens M, Lematty T, Borckardt JJ, Hanlon CA, et al. Repetitive transcranial magnetic stimulation of the dorsolateral prefrontal cortex reduces nicotine cue craving. Biol Psychiatry. 2013 Apr 15;73(8):714–20.

12. Sheffer CE, Mennemeier M, Landes RD, Bickel WK, Brackman S, Dornhoffer J, et al. Neuromodulation of delay discounting, the reflection effect, and cigarette consumption. J Subst Abuse Treat. 2013 Aug;45(2):206–14.

13. Dieler AC, Dresler T, Joachim K, Deckert J, Herrmann MJ, Fallgatter AJ. Can intermittent theta burst stimulation as add-on to psychotherapy improve nicotine abstinence? Results from a pilot study. Eur Addict Res. 2014;20(5):248–53.

14. Dinur-Klein L, Dannon P, Hadar A, Rosenberg O, Roth Y, Kotler M, et al. Smoking cessation induced by deep repetitive transcranial magnetic stimulation of the prefrontal and insular cortices: a prospective, randomized controlled trial. Biol Psychiatry. 2014 Nov 1;76(9):742–9.

15. Prikryl R, Ustohal L, Kucerova HP, Kasparek T, Jarkovsky J, Hublova V, et al. Repetitive transcranial magnetic stimulation reduces cigarette consumption in schizophrenia patients. Prog Neuropsychopharmacol Biol Psychiatry. 2014 Mar 3;49:30–5.

16. Pripfl J, Tomova L, Riecansky I, Lamm C. Transcranial magnetic stimulation of the left dorsolateral prefrontal cortex decreases cue-induced nicotine craving and EEG delta power. Brain Stimul. 2014;7(2):226–33.

17. Ceccanti M, Inghilleri M, Attilia ML, Raccah R, Fiore M, Zangen A, et al. Deep TMS on alcoholics: effects on cortisolemia and dopamine pathway modulation. A pilot study. Can J Physiol Pharmacol. 2015 Apr;93(4):283–90.

18. Girardi P, Rapinesi C, Chiarotti F, Kotzalidis GD, Piacentino D, Serata D, et al. Add-on deep transcranial magnetic stimulation (dTMS) in patients with dysthymic disorder comorbid with alcohol use disorder: a comparison with standard treatment. World J Biol Psychiatry. 2015 Jan;16(1):66–73.

19. Herremans SC, Van Schuerbeek P, De Raedt R, Matthys F, Buyl R, De Mey J, et al. The Impact of Accelerated Right Prefrontal High-Frequency Repetitive Transcranial Magnetic Stimulation (rTMS) on Cue-Reactivity: An fMRI Study on Craving in Recently Detoxified Alcohol-Dependent Patients. PLoS One. 2015;10(8):e0136182.

20. Jm J, G van W, W van den B, Ae G. Resting state connectivity in alcohol dependent patients and the effect of repetitive transcranial magnetic stimulation. European neuropsychopharmacology : the journal of the European College of Neuropsychopharmacology [Internet]. 2015 Dec [cited 2025 Sept 15];25(12). Available from: https://pubmed.ncbi.nlm.nih.gov/26481907/

21. Mishra BR, Praharaj SK, Katshu MZUH, Sarkar S, Nizamie SH. Comparison of anticraving efficacy of right and left repetitive transcranial magnetic stimulation in alcohol dependence: a randomized double-blind study. J Neuropsychiatry Clin Neurosci. 2015;27(1):e54–59.

22. Rapinesi C, Curto M, Kotzalidis GD, Del Casale A, Serata D, Ferri VR, et al. Antidepressant effectiveness of deep Transcranial Magnetic Stimulation (dTMS) in patients with Major Depressive Disorder (MDD) with or without Alcohol Use Disorders (AUDs): a 6-month, open label, follow-up study. J Affect Disord. 2015 Mar 15;174:57–63.

23. Trojak B, Meille V, Achab S, Lalanne L, Poquet H, Ponavoy E, et al. Transcranial Magnetic Stimulation Combined With Nicotine Replacement Therapy for Smoking Cessation: A Randomized Controlled Trial. Brain Stimul. 2015;8(6):1168–74.

24. Bolloni C, Panella R, Pedetti M, Frascella AG, Gambelunghe C, Piccoli T, et al. Bilateral Transcranial Magnetic Stimulation of the Prefrontal Cortex Reduces Cocaine Intake: A Pilot Study. Front Psychiatry. 2016;7:133.

25. Del Felice A, Bellamoli E, Formaggio E, Manganotti P, Masiero S, Cuoghi G, et al. Neurophysiological, psychological and behavioural correlates of rTMS treatment in alcohol dependence. Drug Alcohol Depend. 2016 Jan 1;158:147–53.

26. Hanlon CA, Dowdle LT, Moss H, Canterberry M, George MS. Mobilization of Medial and Lateral Frontal-Striatal Circuits in Cocaine Users and Controls: An Interleaved TMS/BOLD Functional Connectivity Study. Neuropsychopharmacology. 2016 Dec;41(13):3032–41.

27. Herremans SC, De Raedt R, Van Schuerbeek P, Marinazzo D, Matthys F, De Mey J, et al. Accelerated HF-rTMS Protocol has a Rate-Dependent Effect on dACC Activation in Alcohol-Dependent Patients: An Open-Label Feasibility Study. Alcohol Clin Exp Res. 2016 Jan;40(1):196–205.

28. Huang W, Shen F, Zhang J, Xing B. Effect of Repetitive Transcranial Magnetic Stimulation on Cigarette Smoking in Patients with Schizophrenia. Shanghai Arch Psychiatry. 2016 Dec 25;28(6):309–17.

29. Mishra BR, Maiti R, Nizamie SH. Cerebral Hemodynamics With rTMS in Alcohol Dependence: A Randomized, Sham-Controlled Study. J Neuropsychiatry Clin Neurosci. 2016;28(4):319–24.

30. Qiao J, Jin G, Lei L, Wang L, Du Y, Wang X. The positive effects of high-frequency right dorsolateral prefrontal cortex repetitive transcranial magnetic stimulation on memory, correlated with increases in brain metabolites detected by proton magnetic resonance spectroscopy in recently detoxified alcohol-dependent patients. Neuropsychiatr Dis Treat. 2016;12:2273–8.

31. Shen Y, Cao X, Tan T, Shan C, Wang Y, Pan J, et al. 10-Hz Repetitive Transcranial Magnetic Stimulation of the Left Dorsolateral Prefrontal Cortex Reduces Heroin Cue Craving in Long-Term Addicts. Biol Psychiatry. 2016 Aug 1;80(3):e13–14.

32. Terraneo A, Leggio L, Saladini M, Ermani M, Bonci A, Gallimberti L. Transcranial magnetic stimulation of dorsolateral prefrontal cortex reduces cocaine use: A pilot study. Eur Neuropsychopharmacol. 2016 Jan;26(1):37–44.

33. Addolorato G, Antonelli M, Cocciolillo F, Vassallo GA, Tarli C, Sestito L, et al. Deep Transcranial Magnetic Stimulation of the Dorsolateral Prefrontal Cortex in Alcohol Use Disorder Patients: Effects on Dopamine Transporter Availability and Alcohol Intake. Eur Neuropsychopharmacol. 2017 May;27(5):450–61.

34. Baker TE, Lesperance P, Tucholka A, Potvin S, Larcher K, Zhang Y, et al. Reversing the Atypical Valuation of Drug and Nondrug Rewards in Smokers Using Multimodal Neuroimaging. Biol Psychiatry. 2017 Dec 1;82(11):819–27.

35. Hanlon CA, Dowdle LT, Correia B, Mithoefer O, Kearney-Ramos T, Lench D, et al. Left frontal pole theta burst stimulation decreases orbitofrontal and insula activity in cocaine users and alcohol users. Drug Alcohol Depend. 2017 Sept 1;178:310–7.

36. Li X, Du L, Sahlem GL, Badran BW, Henderson S, George MS. Repetitive transcranial magnetic stimulation (rTMS) of the dorsolateral prefrontal cortex reduces resting-state insula activity and modulates functional connectivity of the orbitofrontal cortex in cigarette smokers. Drug Alcohol Depend. 2017 May 1;174:98–105.

37. Li X, Sahlem GL, Badran BW, McTeague LM, Hanlon CA, Hartwell KJ, et al. Transcranial magnetic stimulation of the dorsal lateral prefrontal cortex inhibits medial orbitofrontal activity in smokers. Am J Addict. 2017 Dec;26(8):788–94.

38. Liu Q, Shen Y, Cao X, Li Y, Chen Y, Yang W, et al. Either at left or right, both high and low frequency rTMS of dorsolateral prefrontal cortex decreases cue induced craving for methamphetamine. Am J Addict. 2017 Dec;26(8):776–9.

39. Sahlem GL, Baker NL, George MS, Malcolm RJ, McRae-Clark AL. Repetitive transcranial magnetic stimulation (rTMS) administration to heavy cannabis users. Am J Drug Alcohol Abuse. 2018;44(1):47–55.

40. Su H, Zhong N, Gan H, Wang J, Han H, Chen T, et al. High frequency repetitive transcranial magnetic stimulation of the left dorsolateral prefrontal cortex for methamphetamine use disorders: A randomised clinical trial. Drug Alcohol Depend. 2017 June 1;175:84–91.

41. Kamp D, Engelke C, Wobrock T, Kunze B, Wölwer W, Winterer G, et al. Letter to the Editor: Influence of rTMS on smoking in patients with schizophrenia. Schizophr Res. 2018 Feb;192:481–4.

42. Kearney-Ramos TE, Dowdle LT, Lench DH, Mithoefer OJ, Devries WH, George MS, et al. Transdiagnostic Effects of Ventromedial Prefrontal Cortex Transcranial Magnetic Stimulation on Cue Reactivity. Biol Psychiatry Cogn Neurosci Neuroimaging. 2018 July;3(7):599–609.

43. Kozak K, Sharif-Razi M, Morozova M, Gaudette EV, Barr MS, Daskalakis ZJ, et al. Effects of short-term, high-frequency repetitive transcranial magnetic stimulation to bilateral dorsolateral prefrontal cortex on smoking behavior and cognition in patients with schizophrenia and non-psychiatric controls. Schizophr Res. 2018 July;197:441–3.

44. Liang Q, Lin J, Yang J, Li X, Chen Y, Meng X, et al. Intervention Effect of Repetitive TMS on Behavioral Adjustment After Error Commission in Long-Term Methamphetamine Addicts: Evidence From a Two-Choice Oddball Task. Neuroscience Bulletin. 2018 Jan 16;34.

45. Martinez D, Urban N, Grassetti A, Chang D, Hu MC, Zangen A, et al. Transcranial Magnetic Stimulation of Medial Prefrontal and Cingulate Cortices Reduces Cocaine Self-Administration: A Pilot Study. Front Psychiatry. 2018;9:80.

46. Sheffer CE, Bickel WK, Brandon TH, Franck CT, Deen D, Panissidi L, et al. Preventing relapse to smoking with transcranial magnetic stimulation: Feasibility and potential efficacy. Drug Alcohol Depend. 2018 Jan 1;182:8–18.

47. Zhang L, Cao X, Liang Q, Li X, Yang J, Yuan J. High-frequency repetitive transcranial magnetic stimulation of the left dorsolateral prefrontal cortex restores attention bias to negative information in methamphetamine addicts. Psychiatry Res. 2018 July;265:151–60.

48. Liang Y, Wang L, Yuan TF. Targeting Withdrawal Symptoms in Men Addicted to Methamphetamine With Transcranial Magnetic Stimulation: A Randomized Clinical Trial. JAMA Psychiatry. 2018 Nov 1;75(11):1199–201.

49. McNeill A, Monk RL, Qureshi AW, Makris S, Heim D. Continuous Theta Burst Transcranial Magnetic Stimulation of the Right Dorsolateral Prefrontal Cortex Impairs Inhibitory Control and Increases Alcohol Consumption. Cogn Affect Behav Neurosci. 2018 Dec;18(6):1198–206.

50. Wu GR, Baeken C, Van Schuerbeek P, De Mey J, Bi M, Herremans SC. Accelerated repetitive transcranial magnetic stimulation does not influence grey matter volumes in regions related to alcohol relapse: An open-label exploratory study. Drug Alcohol Depend. 2018 Oct 1;191:210–4.

51. Hanlon CA, Lench DH, Dowdle LT, Ramos TK. Neural Architecture Influences Repetitive Transcranial Magnetic Stimulation-Induced Functional Change: A Diffusion Tensor Imaging and Functional Magnetic Resonance Imaging Study of Cue-Reactivity Modulation in Alcohol Users. Clin Pharmacol Ther. 2019 Oct;106(4):702–5.

52. Jansen JM, van den Heuvel OA, van der Werf YD, de Wit SJ, Veltman DJ, van den Brink W, et al. The Effect of High-Frequency Repetitive Transcranial Magnetic Stimulation on Emotion Processing, Reappraisal, and Craving in Alcohol Use Disorder Patients and Healthy Controls: A Functional Magnetic Resonance Imaging Study. Front Psychiatry. 2019;10:272.

53. Kearney-Ramos TE, Dowdle LT, Mithoefer OJ, Devries W, George MS, Hanlon CA. State-Dependent Effects of Ventromedial Prefrontal Cortex Continuous Thetaburst Stimulation on Cocaine Cue Reactivity in Chronic Cocaine Users. Front Psychiatry. 2019;10:317.

54. Lin J, Liu X, Li H, Yu L, Shen M, Lou Y, et al. Chronic repetitive transcranial magnetic stimulation (rTMS) on sleeping quality and mood status in drug dependent male inpatients during abstinence. Sleep Med. 2019 June;58:7–12.

55. T L, Y L, Y S, X L, Tf Y. Gender does not matter: Add-on repetitive transcranial magnetic stimulation treatment for female methamphetamine dependents. Progress in neuro-psychopharmacology & biological psychiatry [Internet]. 2019 June 8 [cited 2025 Sept 15];92. Available from: https://pubmed.ncbi.nlm.nih.gov/30605708/

56. Prashad S, Dedrick ES, To WT, Vanneste S, Filbey FM. Testing the role of the posterior cingulate cortex in processing salient stimuli in cannabis users: an rTMS study. Eur J Neurosci. 2019 Aug;50(3):2357–69.

57. Sanna A, Fattore L, Badas P, Corona G, Cocco V, Diana M. Intermittent Theta Burst Stimulation of the Prefrontal Cortex in Cocaine Use Disorder: A Pilot Study. Front Neurosci. 2019;13:765.

58. Schluter RS, van Holst RJ, Goudriaan AE. Effects of Ten Sessions of High Frequency Repetitive Transcranial Magnetic Stimulation (HF-rTMS) Add-on Treatment on Impulsivity in Alcohol Use Disorder. Front Neurosci. 2019;13:1257.

59. Biernacki K, Lin MH, Baker TE. Recovery of reward function in problematic substance users using a combination of robotics, electrophysiology, and TMS. Int J Psychophysiol. 2020 Dec;158:288–98.

60. Chen T, Su H, Li R, Jiang H, Li X, Wu Q, et al. The exploration of optimized protocol for repetitive transcranial magnetic stimulation in the treatment of methamphetamine use disorder: A randomized sham-controlled study. EBioMedicine. 2020 Oct;60:103027.

61. Chen T, Su H, Jiang H, Li X, Zhong N, Du J, et al. Cognitive and emotional predictors of real versus sham repetitive transcranial magnetic stimulation treatment response in methamphetamine use disorder. J Psychiatr Res. 2020 July;126:73–80.

62. Gómez Pérez LJ, Cardullo S, Cellini N, Sarlo M, Monteanni T, Bonci A, et al. Sleep quality improves during treatment with repetitive transcranial magnetic stimulation (rTMS) in patients with cocaine use disorder: a retrospective observational study. BMC Psychiatry. 2020 Apr 6;20(1):153.

63. Li X, Hartwell KJ, Henderson S, Badran BW, Brady KT, George MS. Two weeks of image-guided left dorsolateral prefrontal cortex repetitive transcranial magnetic stimulation improves smoking cessation: A double-blind, sham-controlled, randomized clinical trial. Brain Stimul. 2020;13(5):1271–9.

64. The effects of repetitive transcranial magnetic stimulation on cue-induced craving in male patients with heroin use disorder. EBioMedicine. 2020 June 1;56:102809.

65. Liu X, Zhao X, Shen Y, Liu T, Liu Q, Tang L, et al. The effects of DLPFC-targeted repetitive transcranial magnetic stimulation on craving in male methamphetamine patients. Clin Transl Med. 2020 June;10(2):e48.

66. Newman-Norlund RD, Gibson M, McConnell PA, Froeliger B. Dissociable Effects of Theta-Burst Repeated Transcranial Magnetic Stimulation to the Inferior Frontal Gyrus on Inhibitory Control in Nicotine Addiction. Front Psychiatry. 2020;11:260.

67. Perini I, Kämpe R, Arlestig T, Karlsson H, Löfberg A, Pietrzak M, et al. Repetitive transcranial magnetic stimulation targeting the insular cortex for reduction of heavy drinking in treatment-seeking alcohol-dependent subjects: a randomized controlled trial. Neuropsychopharmacology. 2020 Apr;45(5):842–50.

68. Raikwar S, Divinakumar KJ, Prakash J, Khan SA, GuruPrakash KV, Batham S. A sham-controlled trial of repetitive transcranial magnetic stimulation over left dorsolateral prefrontal cortex and its effects on craving in patients with alcohol dependence. Ind Psychiatry J. 2020;29(2):245–50.

69. Su H, Liu Y, Yin D, Chen T, Li X, Zhong N, et al. Neuroplastic changes in resting-state functional connectivity after rTMS intervention for methamphetamine craving. Neuropharmacology. 2020 Sept 15;175:108177.

70. Su H, Chen T, Zhong N, Jiang H, Du J, Xiao K, et al. γ-aminobutyric acid and glutamate/glutamine alterations of the left prefrontal cortex in individuals with methamphetamine use disorder: a combined transcranial magnetic stimulation-magnetic resonance spectroscopy study. Ann Transl Med. 2020 Mar;8(6):347.

71. Su H, Chen T, Jiang H, Zhong N, Du J, Xiao K, et al. Intermittent theta burst transcranial magnetic stimulation for methamphetamine addiction: A randomized clinical trial. Eur Neuropsychopharmacol. 2020 Feb;31:158–61.

72. Yuan J, Liu W, Liang Q, Cao X, Lucas MV, Yuan TF. Effect of Low-Frequency Repetitive Transcranial Magnetic Stimulation on Impulse Inhibition in Abstinent Patients With Methamphetamine Addiction: A Randomized Clinical Trial. JAMA Netw Open. 2020 Mar 2;3(3):e200910.

73. Zhao D, Li Y, Liu T, Voon V, Yuan TF. Twice-Daily Theta Burst Stimulation of the Dorsolateral Prefrontal Cortex Reduces Methamphetamine Craving: A Pilot Study. Front Neurosci. 2020;14:208.

74. Abdelrahman AA, Noaman M, Fawzy M, Moheb A, Karim AA, Khedr EM. A double-blind randomized clinical trial of high frequency rTMS over the DLPFC on nicotine dependence, anxiety and depression. Sci Rep. 2021 Jan 15;11(1):1640.

75. Bozzay ML, Brigido S, van ’t Wout-Frank M, Aiken E, Swift R, Philip NS. Intermittent Theta Burst Stimulation in Veterans with Mild Alcohol Use Disorder. J Affect Disord. 2021 Oct 1;293:314–9.

76. Cardullo S, Gómez Pérez LJ, Cuppone D, Sarlo M, Cellini N, Terraneo A, et al. A Retrospective Comparative Study in Patients With Cocaine Use Disorder Comorbid With Attention Deficit Hyperactivity Disorder Undergoing an rTMS Protocol Treatment. Front Psychiatry. 2021;12:659527.

77. Chen T, Su H, Wang L, Li X, Wu Q, Zhong N, et al. Modulation of Methamphetamine-Related Attention Bias by Intermittent Theta-Burst Stimulation on Left Dorsolateral Prefrontal Cortex. Front Cell Dev Biol. 2021;9:667476.

78. Garza-Villarreal EA, Alcala-Lozano R, Fernandez-Lozano S, Morelos-Santana E, Dávalos A, Villicaña V, et al. Clinical and Functional Connectivity Outcomes of 5-Hz Repetitive Transcranial Magnetic Stimulation as an Add-on Treatment in Cocaine Use Disorder: A Double-Blind Randomized Controlled Trial. Biol Psychiatry Cogn Neurosci Neuroimaging. 2021 July;6(7):745–57.

79. Gupta AK, Kumar A, Chandrashekhar N. Adjuvant treatment with repetitive transcranial magnetic stimulation in freshly diagnosed alcohol-dependence syndrome patients from an industry: An outcome study. Ind Psychiatry J. 2021 Oct;30(Suppl 1):S93–6.

80. Li X, Song GF, Yu JN, Ai SH, Ji Q, Peng Y, et al. Effectiveness and safety of repetitive transcranial magnetic stimulation for the treatment of morphine dependence: A retrospective study. Medicine (Baltimore). 2021 Apr 9;100(14):e25208.

81. Lolli F, Salimova M, Scarpino M, Lanzo G, Cossu C, Bastianelli M, et al. A randomised, double-blind, sham-controlled study of left prefrontal cortex 15 Hz repetitive transcranial magnetic stimulation in cocaine consumption and craving. PLoS One. 2021;16(11):e0259860.

82. Tsai TY, Wang TY, Liu YC, Lee PW, Chang WH, Lu TH, et al. Add-on repetitive transcranial magnetic stimulation in patients with opioid use disorder undergoing methadone maintenance therapy. Am J Drug Alcohol Abuse. 2021 May 4;47(3):330–43.

83. Wang LJ, Mu LL, Ren ZX, Tang HJ, Wei YD, Wang WJ, et al. Predictive Role of Executive Function in the Efficacy of Intermittent Theta Burst Transcranial Magnetic Stimulation Modalities for Treating Methamphetamine Use Disorder-A Randomized Clinical Trial. Front Psychiatry. 2021;12:774192.

84. Zangen A, Moshe H, Martinez D, Barnea-Ygael N, Vapnik T, Bystritsky A, et al. Repetitive transcranial magnetic stimulation for smoking cessation: a pivotal multicenter double-blind randomized controlled trial. World Psychiatry. 2021 Oct;20(3):397–404.

85. Ankit A, Das B, Dey P, Kshitiz KK, Khess CRJ. Efficacy of continuous theta burst stimulation - repetitive trancranial magnetic stimulation on the orbito frontal cortex as an adjunct to naltrexone in patients of opioid use disorder and its correlation with serum BDNF levels: a sham-controlled study. J Addict Dis. 2022;40(3):373–81.

86. Belgers M, Van Eijndhoven P, Markus W, Schene AH, Schellekens A. rTMS Reduces Craving and Alcohol Use in Patients with Alcohol Use Disorder: Results of a Randomized, Sham-Controlled Clinical Trial. J Clin Med. 2022 Feb 11;11(4):951.

87. Bidzinski KK, Lowe DJE, Sanches M, Sorkhou M, Boileau I, Kiang M, et al. Investigating repetitive transcranial magnetic stimulation on cannabis use and cognition in people with schizophrenia. Schizophrenia (Heidelb). 2022 Feb 24;8(1):2.

88. Feng Z, Wu Q, Wu L, Zeng T, Yuan J, Wang X, et al. Effect of High-Frequency Repetitive Transcranial Magnetic Stimulation on Visual Selective Attention in Male Patients With Alcohol Use Disorder After the Acute Withdrawal. Front Psychiatry. 2022;13:869014.

89. Harel M, Perini I, Kämpe R, Alyagon U, Shalev H, Besser I, et al. Repetitive Transcranial Magnetic Stimulation in Alcohol Dependence: A Randomized, Double-Blind, Sham-Controlled Proof-of-Concept Trial Targeting the Medial Prefrontal and Anterior Cingulate Cortices. Biol Psychiatry. 2022 June 15;91(12):1061–9.

90. Jin L, Yuan M, Zhang W, Su H, Wang F, Zhu J, et al. Repetitive transcranial magnetic stimulation modulates coupling among large-scale brain networks in heroin-dependent individuals: A randomized resting-state functional magnetic resonance imaging study. Addict Biol. 2022 Mar;27(2):e13121.

91. Liu Q, Sun H, Hu Y, Wang Q, Zhao Z, Dong D, et al. Intermittent Theta Burst Stimulation vs. High-Frequency Repetitive Transcranial Magnetic Stimulation in the Treatment of Methamphetamine Patients. Front Psychiatry. 2022;13:842947.

92. Marques RC, Marques D, Vieira L, Cantilino A. Left frontal pole repetitive transcranial magnetic stimulation reduces cigarette cue-reactivity in correlation with verbal memory performance. Drug Alcohol Depend. 2022 June 1;235:109450.

93. Martinotti G, Pettorruso M, Montemitro C, Spagnolo PA, Acuti Martellucci C, Di Carlo F, et al. Repetitive transcranial magnetic stimulation in treatment-seeking subjects with cocaine use disorder: A randomized, double-blind, sham-controlled trial. Prog Neuropsychopharmacol Biol Psychiatry. 2022 June 8;116:110513.

94. McNeill AM, Monk RL, Qureshi AW, Makris S, Cazzato V, Heim D. Elevated ad libitum alcohol consumption following continuous theta burst stimulation to the left-dorsolateral prefrontal cortex is partially mediated by changes in craving. Cogn Affect Behav Neurosci. 2022 Feb;22(1):160–70.

95. Mikellides G, Michael P, Psalta L, Stefani A, Schuhmann T, Sack AT. Accelerated Intermittent Theta Burst Stimulation in Smoking Cessation: Placebo Effects Equal to Active Stimulation When Using Advanced Placebo Coil Technology. Front Psychiatry. 2022;13:892075.

96. Moeller SJ, Gil R, Weinstein JJ, Baumvoll T, Wengler K, Fallon N, et al. Deep rTMS of the insula and prefrontal cortex in smokers with schizophrenia: Proof-of-concept study. Schizophr. 2022 Feb 25;8(1):1–9.

97. Shevorykin A, Carl E, Mahoney MC, Hanlon CA, Liskiewicz A, Rivard C, et al. Transcranial Magnetic Stimulation for Long-Term Smoking Cessation: Preliminary Examination of Delay Discounting as a Therapeutic Target and the Effects of Intensity and Duration. Front Hum Neurosci. 2022;16:920383.

98. H S, P Y, T C, D D, N Z, H J, et al. Metabolomics changes after rTMS intervention reveal potential peripheral biomarkers in methamphetamine dependence. European neuropsychopharmacology : the journal of the European College of Neuropsychopharmacology [Internet]. 2022 Mar [cited 2025 Sept 15];56. Available from: https://pubmed.ncbi.nlm.nih.gov/34990999/

99. Zhang T, Song B, Li Y, Duan R, Gong Z, Jing L, et al. Neurofilament Light Chain as a Biomarker for Monitoring the Efficacy of Transcranial Magnetic Stimulation on Alcohol Use Disorder. Front Behav Neurosci. 2022;16:831901.

100. Li X, Toll BA, Carpenter MJ, Nietert PJ, Dancy M, George MS. Repetitive Transcranial Magnetic Stimulation for Tobacco Treatment in Cancer Patients: A Preliminary Report of a One-Week Treatment. J Smok Cessat. 2022;2022:2617146.

101. Liu Q, Xu X, Cui H, Zhang L, Zhao Z, Dong D, et al. High-frequency repetitive transcranial magnetic stimulation of the left dorsolateral prefrontal cortex may reduce impulsivity in patients with methamphetamine use disorders: A pilot study. Front Hum Neurosci. 2022;16:858465.

102. Hu X, Zhang T, Ma H, Zhou X, Wang H, Wang X, et al. Repetitive transcranial magnetic stimulation combined with cognitive behavioral therapy treatment in alcohol-dependent patients: A randomized, double-blind sham-controlled multicenter clinical trial. Front Psychiatry. 2022;13:935491.

103. Johnstone S, Lowe DJE, Kozak-Bidzinski K, Sanches M, Castle DJ, Rabin JS, et al. Neurocognitive moderation of repetitive transcranial magnetic stimulation (rTMS) effects on cannabis use in schizophrenia: a preliminary analysis. Schizophrenia (Heidelb). 2022 Nov 17;8(1):99.

104. Kang T, Ding X, Zhao J, Li X, Xie R, Jiang H, et al. Influence of improved behavioral inhibition on decreased cue-induced craving in heroin use disorder: A preliminary intermittent theta burst stimulation study. J Psychiatr Res. 2022 Aug;152:375–83.

105. Lechner WV, Philip NS, Kahler CW, Houben K, Tirrell E, Carpenter LL. Combined Working Memory Training and Transcranial Magnetic Stimulation Demonstrates Low Feasibility and Potentially Worse Outcomes on Delay to Smoking and Cognitive Tasks: A Randomized 2 × 2 Factorial Design Pilot and Feasibility Study. Nicotine Tob Res. 2022 Nov 12;24(12):1871–80.

106. Sun Y, Wang H, Ku Y. Intermittent Theta-Burst Stimulation Increases the Working Memory Capacity of Methamphetamine Addicts. Brain Sci. 2022 Sept 8;12(9):1212.

107. Wang W, Zhu Y, Wang L, Mu L, Zhu L, Ding D, et al. High-frequency repetitive transcranial magnetic stimulation of the left dorsolateral prefrontal cortex reduces drug craving and improves decision-making ability in methamphetamine use disorder. Psychiatry Res. 2022 Nov;317:114904.

108. McCalley DM, Kaur N, Wolf JP, Contreras IE, Book SW, Smith JP, et al. Medial Prefrontal Cortex Theta Burst Stimulation Improves Treatment Outcomes in Alcohol Use Disorder: A Double-Blind, Sham-Controlled Neuroimaging Study. Biol Psychiatry Glob Open Sci. 2023 Apr;3(2):301–10.

109. Mikellides G, Michael P, Psalta L, Stefani A, Schuhmann T, Sack AT. Accelerated intermittent theta burst stimulation in smoking cessation: No differences between active and placebo stimulation when using advanced placebo coil technology. A double-blind follow-up study. Int J Clin Health Psychol. 2023;23(2):100351.

110. Jin L, Yuan M, Zhang W, Wang L, Chen J, Wang F, et al. Default mode network mechanisms of repeated transcranial magnetic stimulation in heroin addiction. Brain Imaging Behav. 2023 Feb;17(1):54–65.

111. Hoven M, Schluter RS, Schellekens AF, van Holst RJ, Goudriaan AE. Effects of 10 add-on HF-rTMS treatment sessions on alcohol use and craving among detoxified inpatients with alcohol use disorder: a randomized sham-controlled clinical trial. Addiction. 2023 Jan;118(1):71–85.

112. Dong L, Chen WC, Su H, Wang ML, Du C, Jiang XR, et al. Intermittent theta burst stimulation to the left dorsolateral prefrontal cortex improves cognitive function in polydrug use disorder patients: a randomized controlled trial. Front Psychiatry. 2023;14:1156149.

113. Zhang Y, Ku Y, Sun J, Daskalakis ZJ, Yuan TF. Intermittent theta burst stimulation to the left dorsolateral prefrontal cortex improves working memory of subjects with methamphetamine use disorder. Psychol Med. 2023 Apr;53(6):2427–36.

114. Gersner R, Barnea-Ygael N, Tendler A. Moderators of the response to deep TMS for smoking addiction. Front Psychiatry. 2023 Jan 9;13:1079138.

115. 115. Shevorykin A, Carl E, Liskiewicz A, Hanlon CA, Bickel WK, Mahoney MC, et al. Perceived research burden of a novel therapeutic intervention: A study of transcranial magnetic stimulation for smoking cessation. Front Rehabil Sci [Internet]. 2023 Mar 3 [cited 2025 Sept 15];4. Available from: https://www.frontiersin.org/journals/rehabilitation-sciences/articles/10.3389/fresc.2023.1054456/full

116. Upton S, Brown AA, Ithman M, Newman-Norlund R, Sahlem G, Prisciandaro JJ, et al. Effects of Hyperdirect Pathway Theta Burst Transcranial Magnetic Stimulation on Inhibitory Control, Craving, and Smoking in Adults With Nicotine Dependence: A Double-Blind, Randomized Crossover Trial. Biol Psychiatry Cogn Neurosci Neuroimaging. 2023 Nov;8(11):1156–65.

117. Ibrahim C, Tang VM, Blumberger DM, Malik S, Tyndale RF, Trevizol AP, et al. Efficacy of insula deep repetitive transcranial magnetic stimulation combined with varenicline for smoking cessation: A randomized, double-blind, sham controlled trial. Brain Stimul. 2023;16(5):1501–9.

118. Gong H, Huang Y, Zhu X, Lu W, Cai Z, Zhu N, et al. Impact of combination of intermittent theta burst stimulation and methadone maintenance treatment in individuals with opioid use disorder: A comparative study. Psychiatry Res. 2023 Sept;327:115411.

119. Gerace E, Baldi S, Salimova M, Di Gloria L, Curini L, Cimino V, et al. Oral and fecal microbiota perturbance in cocaine users: Can rTMS-induced cocaine abstinence support eubiosis restoration? iScience. 2023 May 19;26(5):106627.

120. Upton S, Brown AA, Golzy M, Garland EL, Froeliger B. Right inferior frontal gyrus theta-burst stimulation reduces smoking behaviors and strengthens fronto-striatal-limbic resting-state functional connectivity: a randomized crossover trial. Front Psychiatry [Internet]. 2023 June 28 [cited 2025 Sept 15];14. Available from: https://www.frontiersin.org/journals/psychiatry/articles/10.3389/fpsyt.2023.1166912/full

121. Ding X, Li X, Xu M, He Z, Jiang H. The effect of repetitive transcranial magnetic stimulation on electroencephalography microstates of patients with heroin-addiction. Psychiatry Res Neuroimaging. 2023 Mar;329:111594.

122. Zhao K, Fonzo GA, Xie H, Oathes DJ, Keller CJ, Carlisle NB, et al. Discriminative functional connectivity signature of cocaine use disorder links to rTMS treatment response. Nat Mental Health. 2024 Apr;2(4):388–400.

123. Li X, Caulfield KA, Hartwell KJ, Henderson S, Brady KT, George MS. Reduced executive and reward connectivity is associated with smoking cessation response to repetitive transcranial magnetic stimulation: A double-blind, randomized, sham-controlled trial. Brain Imaging Behav. 2024 Feb;18(1):207–19.

124. Selim MK, Harel M, De Santis S, Perini I, Sommer WH, Heilig M, et al. Repetitive deep TMS in alcohol dependent patients halts progression of white matter changes in early abstinence. Psychiatry Clin Neurosci. 2024 Mar;78(3):176–85.

125. Cardullo S, Gómez Pérez LJ, Terraneo A, Gallimberti L, Mioni G. Time perception in stimulant-dependent participants undergoing repetitive transcranial magnetic stimulation. Behav Brain Res. 2024 Mar 5;460:114816.

126. Jordan T, Apostol MR, Nomi J, Petersen N. Unraveling Neural Complexity: Exploring Brain Entropy to Yield Mechanistic Insight in Neuromodulation Therapies for Tobacco Use Disorder. bioRxiv. 2023 Sept 13;2023.09.12.557465.

127. Addicott MA, Kinney KR, Saldana S, Ip EHS, DeMaioNewton H, Bickel WK, et al. A randomized controlled trial of intermittent theta burst stimulation to the medial prefrontal cortex for tobacco use disorder: Clinical efficacy and safety. Drug Alcohol Depend. 2024 May 1;258:111278.

128. Bellini BB, Scholz JR, Abe TO, Arnaut D, Tonstad S, Alberto RL, et al. Does deep TMS really works for smoking cessation? A prospective, double blind, randomized, sham controlled study. Prog Neuropsychopharmacol Biol Psychiatry. 2024 June 8;132:110997.

129. Du X, Choa FS, Chiappelli J, Bruce H, Kvarta M, Summerfelt A, et al. Combining neuroimaging and brain stimulation to test alternative causal pathways for nicotine addiction in schizophrenia. Brain Stimul. 2024;17(2):324–32.

130. Li S, Ma X, Chen H, Wang M, Zheng Y, Yang B, et al. rTMS effects on urges and severity of tobacco use disorder operate independently of a retrieval-extinction component and involve frontal-striatal pathways. J Affect Disord. 2024 Mar 15;349:21–31.

131. Li Y, Yang B, Ma J, Gao S, Zeng H, Wang W. Assessment of rTMS treatment effects for methamphetamine use disorder based on EEG microstates. Behav Brain Res. 2024 May 8;465:114959.

132. Padula CB, McCalley DM, Tenekedjieva LT, MacNiven K, Rauch A, Morales JM, et al. A pilot, randomized clinical trial: Left dorsolateral prefrontal cortex intermittent theta burst stimulation improves treatment outcomes in veterans with alcohol use disorder. Alcohol Clin Exp Res (Hoboken). 2024 Jan;48(1):164–77.

133. Rakesh G, Adams TG, Morey RA, Alcorn JL, Khanal R, Su AE, et al. Intermittent theta burst stimulation and functional connectivity in people living with HIV/AIDS who smoke tobacco cigarettes: a preliminary pilot study. Front Psychiatry. 2024;15:1315854.

134. Rasgado-Toledo J, Issa-Garcia V, Alcalá-Lozano R, Garza-Villarreal EA, González-Escamilla G. Cortical and subcortical microstructure integrity changes after repetitive transcranial magnetic stimulation therapy in cocaine use disorder and relates to clinical outcomes. Addict Biol. 2024 Feb;29(2):e13381.

135. Ren Z, Mu L, Wang L, Xia L, Song P, Wang Y, et al. Predictive role of impulsivity, anxiety, and depression in the efficacy of intermittent theta burst transcranial magnetic stimulation modalities for treating methamphetamine use disorder: A randomized clinical trial. J Subst Use Addict Treat. 2024 Jan;156:209189.

136. Li S, Zhang Z, Jiang A, Ma X, Wang M, Ni H, et al. Repetitive transcranial magnetic stimulation reshaped the dynamic reconfiguration of the executive and reward networks in individuals with tobacco use disorder. J Affect Disord. 2024 Nov 15;365:427–36.

137. Wang T, Li R, Chen D, Xie M, Li Z, Mao H, et al. Modulation of High-Frequency rTMS on Reward Circuitry in Individuals with Nicotine Dependence: A Preliminary fMRI Study. Neural Plast. 2024;2024:5673579.

138. LoFaro FM, Jordan T, Apostol MR, Steele VR, Konova AB, Petersen N. Stimulating the posterior parietal cortex reduces self-reported risk-taking propensity in people with tobacco use disorder. Addict Neurosci. 2024 Sept;12:100160.

139. Sahlem GL, Kim B, Baker NL, Wong BL, Caruso MA, Campbell LA, et al. A preliminary randomized controlled trial of repetitive transcranial magnetic stimulation applied to the left dorsolateral prefrontal cortex in treatment seeking participants with cannabis use disorder. Drug Alcohol Depend. 2024 Jan 1;254:111035.

140. Liu Q, Cui H, Li J, Shen Y, Zhang L, Zheng H. Modulation of dlPFC function and decision-making capacity by repetitive transcranial magnetic stimulation in methamphetamine use disorder. Transl Psychiatry. 2024 July 8;14(1):280.

141. Li Y, Yang B, Ma J, Li Y, Zeng H, Zhang J. Assessment of rTMS treatment effects for methamphetamine addiction based on EEG functional connectivity. Cogn Neurodyn. 2024 Oct 1;18(5):2373–86.

